# Characterizing common and rare variations in non-traditional glycemic biomarkers using multivariate approaches on multi-ancestry ARIC study

**DOI:** 10.1101/2023.06.13.23289200

**Authors:** Debashree Ray, Stephanie J. Loomis, Sowmya Venkataraghavan, Adrienne Tin, Bing Yu, Nilanjan Chatterjee, Elizabeth Selvin, Priya Duggal

## Abstract

Glycated hemoglobin, fasting glucose, glycated albumin, and fructosamine are biomarkers that reflect different aspects of the glycemic process. Genetic studies of these glycemic biomarkers can shed light on unknown aspects of type 2 diabetes genetics and biology. While there exists several GWAS of glycated hemoglobin and fasting glucose, very few GWAS have focused on glycated albumin or fructosamine. We performed a multi-phenotype GWAS of glycated albumin and fructosamine from 7,395 White and 2,016 Black participants in the Atherosclerosis Risk in Communities (ARIC) study on the common variants from genotyped/imputed data. We found 2 genome-wide significant loci, one mapping to known type 2 diabetes gene (*ARAP1/STARD10*, *p* = 2.8 × 10^−8^) and another mapping to a novel gene (*UGT1A*, *p* = 1.4 × 10^−8^) using multi-omics gene mapping strategies in diabetes-relevant tissues. We identified additional loci that were ancestry-specific (e.g., *PRKCA* from African ancestry individuals, *p* = 1.7 × 10^−8^) and sex-specific (*TEX29* locus in males only, *p* = 3.0 × 10^−8^). Further, we implemented multi-phenotype gene-burden tests on whole-exome sequence data from 6,590 White and 2,309 Black ARIC participants. Eleven genes across different rare variant aggregation strategies were exome-wide significant only in multi-ancestry analysis. Four out of 11 genes had notable enrichment of rare predicted loss of function variants in African ancestry participants despite smaller sample size. Overall, 8 out of 15 loci/genes were implicated to influence these biomarkers via glycemic pathways. This study illustrates improved locus discovery and potential effector gene discovery by leveraging joint patterns of related biomarkers across entire allele frequency spectrum in multi-ancestry analyses. Most of the loci/genes we identified have not been previously implicated in studies of type 2 diabetes, and future investigation of the loci/genes potentially acting through glycemic pathways may help us better understand risk of developing type 2 diabetes.

## INTRODUCTION

Type 2 diabetes (T2D), characterized by hyperglycemia or elevated blood glucose levels, is a major public health concern worldwide, including in the US. Hyperglycemia is commonly measured by the classical biomarkers, fasting glucose and hemoglobin A1c (HbA1c). However, fasting glucose measurement requires patient preparation (8-hour fasting), exhibits moderate within-person variability, has sample stability issues and is affected by illness and stress, while measurement of HbA1c is costly, requires whole blood, is hemoglobin dependent and may be inaccurate when life of red blood cell is altered.^1, 2^ Non-traditional biomarkers such as fructosamine and glycated albumin show promise as alternatives in the face of these limitations.^1, 3–8^ Fructosamine reflects glucose bound to total serum protein while glycated albumin reflects glucose bound to albumin, the most prevalent serum protein. These biomarkers reflect average blood glucose levels over the previous 2-3 weeks.^9^ They improve risk stratification of diabetes and its long-term complications, provide information complementary to those from classical glycemic biomarkers, and hold promise in providing unique insights into hyperglycemia and diabetes pathophysiology.^10–13^ Non-traditional biomarkers, although not commonly used clinically in the US^14^, have gained traction in their usage around the world. Organizations in India, Australia, and the UK recommend fructosamine measurement in individuals with known erythrocyte conditions where HbA1c testing is unreliable^1^. Glycated albumin is regularly used clinically to monitor short-term changes in glycemic control for diabetes in China, Japan, and South Korea, and has been cleared by the FDA for clinical use in the US.^1, 15^

Genetic studies of traditional measures of hyperglycemia like fasting glucose and HbA1c– have improved our understanding of the genetic mechanisms that may influence hyperglycemia and lead to T2D.^16^ The genetic architecture of these biomarkers is complex. A large multi-ancestry study evaluating fasting glucose and HbA1c in >280,000 individuals without diabetes identified 102 loci for fasting glucose and 127 loci for HbA1c.^17^ However, little is known about the genetics of the non-traditional biomarkers of T2D. Our previous genome-wide association study (GWAS) on fructosamine and glycated albumin identified several associated genetic variants, including a known T2D-related missense variant in *GCKR* and a novel missense variant in *RCN3*, which may impact fructosamine in a non-glycemic manner.^18^ Using SOLAR-Eclipse^19^, the estimated narrow-sense heritability– representing heritability from the additive effects of variants across the entire genome– for both fructosamine and glycated albumin are substantial (*h*^2^ = 0.44 [SE 0.13] and 0.45 [SE 0.13], respectively) and even greater than HbA1c (*h*^2^ = 0.34 [SE 0.13]).^20^ Yet, the SNP-based heritability estimates (*h*^2^)– representing heritability from common variants– using GCTA^21^ are 0.11 (SE 0.03) for fructosamine and 0.10 (SE 0.04) for glycated albumin compared to 0.17 (SE 0.04) for HbA1c.^20^ These estimates suggest that there remain unidentified variants, including possibly rare variants, associated with these biomarkers. Just like GWAS of traditional glycemic biomarkers have improved knowledge of T2D pathophysiology at a time when T2D was studied as a dichotomous trait alone^16^, it is important to understand the genetic basis of these complementary non-traditional glycemic biomarkers that may inform yet-unknown mechanisms modulating glucose control and eventually leading to T2D. Identifying additional variants associated with fructosamine and glycated albumin not found in our previous GWAS^18^ require methods that exploit additional information on these biomarkers and/or genotypes to improve statistical power.

One way to achieve improved power to detect disease-associated variants is through multivariate analysis of underlying disease-related phenotypes.^22–24^ Multivariate phenotype methods jointly analyze two or more phenotypes (e.g., multiple glycemic biomarkers) by accounting for the correlation structure of the phenotypes, and tests if a genetic variant is statistically associated with at least one phenotype. Such approaches can identify pleiotropic variants that are otherwise hard to capture using standard single-phenotype analyses.^24–26^ In addition, they may capture the genetic association of a single phenotype by harnessing joint patterns across related phenotypes.^24^ Bivariate heritability estimates (narrow-sense *h*^2^ = 0.46 using SOLAR-Eclipse, SNP-based 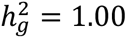 using GCTA)^20^ indicate shared genetics between fructosamine and glycated albumin, and hence bivariate analysis of these phenotypes may help identify genetic variants associated with both these biomarkers.

In this paper, we aimed to expand the current inventory of genetic variants influencing these non-traditional biomarkers of hyperglycemia. We investigated common genetic associations using a data-adaptive multi-phenotype single variant test (metaUSAT^27^) on genotyped/imputed data, and examined rare genetic associations using multi-phenotype gene-based method (GAMuT^28^) on exome sequence data from the Atherosclerosis Risk in Communities (ARIC) Study^29^. We also investigated if genetic findings for these biomarkers differed by genetic ancestry and sex. Finally we examined if the identified common variant loci and genes containing rare variants were potentially acting through glycemic pathways.

## METHODS

### Study population

The ARIC study is a prospective cohort study initiated in 1987 to examine cardiovascular disease risk factors. It follows 15,792 middle-aged adults from four study centers: Jackson, Mississippi; Forsyth county, North Carolina; Washington County, Maryland; and suburban Minneapolis, Minnesota^29^. The baseline visit occurred in 1987-1989 with eight follow-up visits. Informed consent was obtained from study participants, and relevant institutional review boards approved the study protocols. We included study participants who had non-missing genotyping or exome sequencing data. A total of 14,348 individuals attended ARIC visit 2, from whom we excluded participants with diagnosed diabetes (*n* = 1,256), missing diabetes status (*n* = 4), or missing fructosamine or glycated albumin measures (*n* = 929; **Figure S1**). We included 9,411 individuals (*n* = 7,395 self-reported White and 2,016 self-reported Black) who were genotyped and passed quality control, and 8,898 individuals (*n* = 6,589 self-reported White, 2,309 self-reported Black) who were exome sequenced and passed quality control.

### Race, ethnicity, and genetic ancestry

ARIC participants reported self-identified race in a questionnaire at baseline by choosing from the options “Black”, “White”, “Asian”, and “American or Alaskan Indian”. It is important to note that race/ethnicity categories are non-biological, space- and time-dependent social constructs.^30, 31^ Differences in clinical measures and outcomes between racial/ethnic categories do not necessarily indicate biological differences between them, but genetic ancestry determinations are important to avoid population stratification. Thus, we analyzed all individuals together, and performed genetic ancestry-stratified analyses. See **Supplementary Methods** for details on genetic ancestry determination of ARIC participants using 1000 Genomes data and principal component analysis**. Genotyping, imputation, exome sequencing, and quality control**

Samples were genotyped on the Affymetrix 6.0 array and imputed to 1000 Genomes Phase I (March 2012) separately by ancestry using IMPUTE2^32^. Poor quality samples and variants were removed using usual quality control metrics, including removal of first-degree relatives (**Supplementary Methods**). The final dataset included 7,827,582 genotyped/imputed SNPs with MAF ≥ 5%. Blood-based whole exome sequencing of the samples were done on the Illumina HiSeq 2000 or 2500 platform (San Diego, CA), and several quality control measures were implemented (**Supplementary Methods**). The post quality control sample included genetic data on 2,556,859 single nucleotide variants and 76,133 indels from the exome.

### Hyperglycemia biomarkers

Both fructosamine and glycated albumin were measured from serum samples collected at visit 2 (1990-1992). Samples were stored at -70°C. Fructosamine (Roche Diagnostics, Indianapolis IN, USA) and glycated albumin (GA-L Asashi Kasei Pharma Corporation, Tokyo, Japan) were measured in 2012-2013 using a Roche Modular P800 system. In this study, glycated albumin was expressed as percent glycated albumin: [(glycated albumin concentration (g/dL) / serum albumin concentration (g/dL)) ×100/1.14] + 2.9.

### Statistical association analyses

For statistical analyses, we natural log transformed values of both biomarkers to account for skewed distributions. For the multi-ancestry analysis, we obtained biomarker residuals after adjusting each biomarker for age, sex, race-center variable and the top 10 genetic principal components (PCs). While race/ethnicity indicator and at least ten genetic PCs in the model can sufficiently account for admixture^33^, we considered a concatenated race-center variable with 5 distinct categories (Black_Jackson, Black_Forsyth, White_Forsyth, White_Washington, White_Minneapolis)^10, 11^ due to disparate distribution of self-reported race across ARIC study centers. We used the biomarker residuals as outcomes in all genetic association analyses. To evaluate effect of body mass index (BMI) on these glycemic traits, we also considered BMI-adjusted biomarker residuals. All single variant analyses focus on variants with MAF ≥ 5%.

#### Single variant multi-phenotype analysis

To maximize our data usage, we used the data-adaptive multivariate methods that account for the correlations in phenotype structure and the relatedness of phenotypes. Data-adaptive multi-phenotype (multivariate) methods– usually based on GWAS summary statistics– ensure robust power performance across different alternatives^34^ that vary from one variant to the next. We first obtained GWAS summary statistics by performing linear regression analysis of the biomarker residuals on each variant from the genotyped/imputed data using an additive genetic model in PLINK^35^. We then implemented metaUSAT^25, 27^ on the GWAS summary statistics, and tested the joint association of each variant with the two biomarker residuals (details in **Supplementary Methods**).

#### Rare variant set-based multi-phenotype analysis

A single variant test of multiple phenotypes is not ideal for exome sequence data due to the large number of low-frequency and rare variants.^34^ We implemented gene-based multivariate analysis of the biomarkers on variants with MAF< 5% from the exome sequence data using GAMuT^28^, a non-parametric test of no association between a set of phenotypes and a set of genetic variants. Similar to previous human exome sequencing studies^36–38^, we defined 4 different, sometimes nested, sets of functionally important rare and common variants called variant masks using SnpEff^39^: (1) protein-truncating variants (PTVs) at any allele frequency; (2) PTVs in Mask 1 plus missense variants with MAF<5%; (3) pLOFs at any allele frequency; (4) pLOFs with MAF<5%. We also created a mask involving rare deleterious missense variants and rare pLOFs; however, the resultant mask was identical to Mask 4. Gene regions were defined in SnpEff using GRCh37 Ensembl release 75. To assess cumulative effect of variants in a gene, we collapsed all variants into a single burden genotype as done previously^40^. See **Supplementary Methods** for more details.

We used an updated version of GAMuT implementation in MSKAT^41^ R package for more accurate small p-value computation. For the biomarkers, we considered phenotype similarity matrix using both projection and linear kernels (i.e., two different ways of summarizing multivariate phenotype information). For each mask, we only included genes with at least 3 qualifying variants and a burden minor allele count (MAC) of 5 or more^42^. For our multi-ancestry analysis, Masks 1-4 respectively contained 4882, 17726, 1396, and 1375 genes. We used a common, conservative Bonferroni-corrected exome-wide significance threshold of 2.5 × 10^−6^ for all gene-based tests (4 masks × 2 phenotype similarity matrices).

### Stratified analyses

In addition to sex-combined multi-ancestry analyses described above, we performed both sex- and ancestry-stratified analyses. For sex-stratified analysis, we obtained biomarker residuals in males and females separately, which were then used as outcomes in all multi-ancestry models for association analysis. For ancestry-stratified analysis, biomarker residuals and genetic PCs were obtained in each group separately. We only considered BMI-unadjusted models in our stratified analyses since results from the sex-combined multi-ancestry analysis were qualitatively similar with and without BMI adjustment. In our stratified single variant analyses, we additionally explored effect size estimates and their confidence intervals via forest plots since p-values can be confounded by differences in sample sizes.

### Locus annotation

Independent loci from single variant analyses were defined using FUMA (SNP2GENE function, v1.3.7) by clumping all significant SNPs (definitions of statistical significance differ among analyses) in a ±500 Kb radius and with linkage disequilibrium (LD) *r*^2^ > 0.2. We chose 1000G Phase 3 EUR, AFR, and ALL as LD reference panels for European ancestry, African ancestry, and multi-ancestry groups respectively. Lead SNP was defined as the most significant SNP in a locus. In the Results section, for ease of referencing, we annotated identified loci with gene names based on proximity to the lead SNPs regardless of evidence of causal gene, if any, in the literature.

### Plasma protein quantitative trait locus analysis

To characterize the regulatory effects of the significant signals and prioritize functional genes, we searched for overlap between detected genetic determinants from our single variant multi-phenotype analysis and plasma protein quantitative trait loci (pQTLs). We used summary statistics from a recent pQTL study on ARIC cohort that reported results from *cis* genetic regulation of the plasma proteome measured using SomaLogic V4 platform in White (*n* = 7,213) and Black (*n* = 1,871) participants separately.^43^ *Cis*-regions were defined as ±500 Kb of the transcription start site (TSS) of the target protein-coding gene.^43^ From each locus identified in our GWAS, we cross-referenced to see if the lead SNP was a significant *cis*-pQTL at a conservative threshold of 5 × 10^−8^. We also cross-referenced a ±50 Kb radius around our lead SNPs to determine if it contained any significant *cis*-pQTL.

### Functional gene prioritization

We additionally used three strategies in FUMA to perform functional gene mapping of the identified loci: positional mapping, eQTL mapping, and 3D chromatin interaction mapping. We used GTEx v8 expression data^44^ from T2D-relevant tissues (adipose, liver, skeletal muscle, pancreas)^45^, and transcript expression data from human pancreatic islets^46^ for eQTL mapping, and Hi-C data in GSE87112^47^ for 3D chromatin interaction mapping. See **Supplementary Methods** for further details.

### Characterization of detected loci and genes as glycemic vs non-glycemic

#### Loci identified from single variant multi-phenotype analysis

Genetic basis of fructosamine and glycated albumin could be influenced through non-glycemic pathways. To understand biological pathways of these biomarkers that relate to glycemia, we repeated all our analyses by conditioning on fasting glucose and followed a classification algorithm similar to those used previously^48, 49^. We classified a locus as “glycemic” if (a) effect size estimates of its lead SNP for both fructosamine and glycated albumin are reduced by at least 25% ^48^ when fasting glucose is adjusted in the model, thus indicating a glycemic pathway mediated by fasting glucose, or (b) the lead SNP is associated (*p* < 10^−4^) with any glycemic trait except HbA1c (since many genetic associations of HbA1c are often driven through associations with blood cell traits^48^) in a phenome-wide association (PheWAS) analysis of all traits available in Common Metabolic Diseases Knowledge Portal (CMD-KP)^50^ as of July 22, 2022. If association effect sizes were unchanged or increased in the mediation analysis and not associated with any glycemic trait in CMD-KP, we used single-phenotype association results with fasting glucose (*p* < 0.005) in ARIC to classify them as “glycemic”. For the remaining unclassified variants, we marked the lead SNP as “maybe glycemic” if associated with HbA1c in CMD-KP (*p* < 10^−4^) or in ARIC (*p* < 0.005), else marked them as “non-glycemic”. Both fasting glucose and HbA1c measurements were from ARIC visit 2 (1990-1992) and we obtained their single-phenotype association results as before. It is possible for a lead SNP to be associated with both glycemic traits and other non-glycemic traits (e.g., blood-related traits) in CMD-KP; we classified such variants as “glycemic” since in our classification we are primarily concerned about loci affecting fructosamine and glycated albumin in a manner not reflecting ambient glycemia^48^. More details about glycemic traits chosen from CMD-KP and other related choices are provided in **Supplementary Methods**.

#### Genes identified from set-based multi-phenotype analysis

Absence of effect size estimates for the set-based analyses of variants impedes us from quantitatively assessing effect size change in mediation analyses with fasting glucose. Instead, we classified the exome-wide significant genes as “glycemic” if rare variant gene-level associations for the genes were suggestively associated (*p* < 0.005) with any glycemic trait except HbA1c in CMD-KP. We excluded genetic associations with HbA1c from this definition since they are often driven through associations with blood cell traits. The gene-level associations in CMD-KP were based on seven different masks described previously^51^, and were determined as part of the multi-ancestry AMP T2D-GENES exome sequence analysis and the AMP T2D-GENES quantitative trait exome sequence analysis studies. If a gene was not associated with any glycemic trait in CMD-KP, we used single-phenotype set-based GAMuT analysis of fasting glucose (*p* < 0.005) in ARIC to classify the gene as “glycemic”. The remaining unclassified genes were marked as “maybe glycemic” if associated with HbA1c in either CMD-KP or ARIC; otherwise, were marked as “non-glycemic”.

## RESULTS

### Study characteristics

This study included 9,411 multi-ancestry participants (78% European ancestry, 21% African ancestry) with genotyped data and 8,898 multi-ancestry participants (67% European ancestry, 23% African ancestry) with exome sequencing data. Over half were female, and mean age was around 57 years. Values for all glycemic biomarkers were higher among African ancestry individuals than European ancestry (**Table S1**). Correlation between the biomarkers was strong (*r* = 0.79), and estimates were similar in the genotyped and the exome sequenced samples (**Table S2**).

### Locus discovery from multi-phenotype analysis of common variants

At genome-wide significance level 5 × 10^−8^, the joint analysis of fructosamine and glycated albumin using BMI-unadjusted model on sex-combined multi-ancestry data identified 2 loci: the well-known glycemic locus 11q13.4 (*ARAP1*/*STARD10*, rs116714277-T *p* = 2.8 × 10^−8^)^52^ and the locus 17q24.2 (*PRKCA*, rs59443763-C *p* = 1.4 × 10^−8^) implicated in lipids and cardiovascular traits^50, 53^ (**Table 1**). Both the *ARAP1*/*STARD10* and *PRKCA* loci were only identified in African ancestry individuals as the lead SNPs at these loci were removed due to poor quality in our European ancestry sample (**Figure S2**). These lead SNPs rs116714277 and rs59443763 are extremely rare in non-Finnish European population (MAF 0.03% and 0.06% respectively) as compared to the African/African American population (MAF 7.3% and 6.8% respectively).^54^ We found minimal differences in the genome-wide metaUSAT p-values of BMI-unadjusted and BMI-adjusted models (**Figure S3**).

**Table 1.** Association results for the most significant SNPs of the loci identified from multivariate analysis of fructosamine and glycated albumin using BMI-unadjusted model on sex-combined genotyped/imputed data. These loci were identified by metaUSAT at the genome-wide threshold of 5 × 10^−8^ plus an additional locus that just missed this threshold. For a SNP, the metaUSAT p-value < 5 × 10^−8^ indicates its statistically significant association with at least one of fructosamine and glycated albumin at the genome-wide level, which may or may not be significant in single-phenotype analysis. The effect sizes and p-values for individual phenotypes are from BMI-unadjusted model only. All results correspond to the most significant SNP (lead SNP) in a locus.

| Locus | Nearest gene | rsID<br>(lead SNP) | Position<br>(hg19) | Effect<br>allele | Fructosamine |  | Glycated Albumin |  | Multivariate analysis<br>(metaUSAT) |  | *cis*-eQTL mapping in T2D-relevant<br>tissues<sup>#</sup> |
| --- | --- | --- | --- | --- | --- | --- | --- | --- | --- | --- | --- |
| | | | | | Effect<br>size $\beta^{\S}$ | P-value | Effect<br>size $\beta^{\S}$ | P-value | BMI-unadjusted<br>p-value | BMI-adjusted<br>p-value | |
| Multi-ancestry (*n* = 9411) |  |  |  |  |  |  |  |  |  |  |  |
| 11q13.4<sup>¶</sup> | *ARAPI1*/<br>*STARD10* | rs116714277 | 72473447 | T | 0.044 | $2.4 \times 10^{-7}$ | 0.055 | $5.0 \times 10^{-8}$ | $2.8 \times 10^{-8}$ | $3.1 \times 10^{-8}$ | Muscle_skeletal |
| 17q24.2 | *PRKCA* | rs59443763 | 64526988 | C | 0.037 | $1.6 \times 10^{-6}$ | 0.053 | $3.3 \times 10^{-9}$ | $1.4 \times 10^{-8}$ | $6.7 \times 10^{-8}$ | No *cis*-eQTL in this locus |
| European ancestry (*n* = 7359) |  |  |  |  |  |  |  |  |  |  |  |
| 2p23.3 | *GCKR* | rs1260326 | 27730940 | T | -0.0036 | $2.0 \times 10^{-2}$ | -0.010 | $4.0 \times 10^{-8}$ | $5.2 \times 10^{-8}$ | $6.4 \times 10^{-8}$ | Adipose_subcutaneous,<br>Adipose_visceral, Muscle_skeletal,<br>Pancreas |
| 2q37.1<sup>¶</sup> | *UGT1A1*<sup>*</sup> | rs887829 | 234668570 | T | 0.0074 | $4.7 \times 10^{-6}$ | 0.00079 | $6.8 \times 10^{-1}$ | $2.2 \times 10^{-8}$ | $2.1 \times 10^{-8}$ | Adipose_subcutaneous, Liver, Pancreas |
| 19q13.33 | *FCGRT* | rs59774409 | 50016748 | T | -0.018 | $1.6 \times 10^{-7}$ | -0.022 | $1.2 \times 10^{-7}$ | $2.4 \times 10^{-8}$ | $7.2 \times 10^{-7}$ | Adipose_subcutaneous,<br>Adipose_visceral, Liver,<br>Muscle_skeletal, Pancreas |
| African ancestry (*n* = 2004) |  |  |  |  |  |  |  |  |  |  |  |
| 11q13.4<sup>¶</sup> | *ARAPI1*/<br>*STARD10* | rs116714277 | 72473447 | T | 0.045 | $1.5 \times 10^{-7}$ | 0.056 | $2.3 \times 10^{-8}$ | $2.2 \times 10^{-8}$ | $3.2 \times 10^{-8}$ | Muscle_skeletal |
| 11q22.1 | *RP11-115E19.1* | rs2438321 | 98500410 | G | 0.039 | $2.1 \times 10^{-9}$ | 0.033 | $1.2 \times 10^{-5}$ | $1.3 \times 10^{-8}$ | $6.2 \times 10^{-8}$ | No *cis*-eQTL in this locus |
| 17q24.2 | *PRKCA* | rs59443763 | 64526988 | C | 0.037 | $1.1 \times 10^{-6}$ | 0.053 | $3.4 \times 10^{-9}$ | $1.7 \times 10^{-8}$ | $6.5 \times 10^{-8}$ | No *cis*-eQTL in this locus |

<sup>§</sup> $\beta$ denotes mean change in log(outcome) for every additional copy of the effect allele.
<sup>#</sup>The eQTL datasets consist primarily of European ancestry individuals, which may lead to limited eQTL findings for African ancestry participants.
<sup>¶</sup>Novel locus for fructosamine and/or glycated albumin (may or may not be novel for type 2 diabetes).
\*This region includes multiple alternatively spliced genes including *UGT1A1/UGT1A3/UGT1A4/UGT1A5/UGT1A10*.

### Effect size heterogeneity at two loci of common variants between European and African ancestries

The multivariate analysis in African ancestry identified 3 significant loci: 11q13.4 (*ARAP1*/*STARD10*, rs116714277-T *p* = 2.2 × 10^−8^), 11q22.1 (*RP11-115E19.1*, rs2438321-G *p* = 1.3 × 10^−8^), and 17q24.2 (*PRKCA*, rs59443763-C *p* = 1.7 × 10^−8^) (**Table 1**). The locus near *RP11-115E19.1* showed significant effect size heterogeneity at the lead SNP (**Figure S2**), and some allele frequency differences between European (non-Finnish MAF 27.2%) and African/African American (MAF 12.4%) populations^54^. ^50^We did not find any qualitative difference overall in significance when BMI-adjusted models were used (**Figure S4**). Among European ancestry individuals, 2 loci were genome-wide significant: 2q37.1 (*UGT1A* region, rs887829-T *p* = 2.2 × 10^−8^) and 19q13.33 (*FCGRT*, rs59774409-T *p* = 2.4 × 10^−8^). An additional well-known lipids and glycemic locus just missed the GWAS threshold: 2p23.3 (*GCKR*, rs1260326-T *p* = 5.2 × 10^−8^).^55^

The *UGT1A* locus was not significant in the multi-ancestry GWAS of either fructosamine or glycated albumin alone. The *UGT1A* region lead SNP showed some heterogeneity in effect sizes between ancestries (**Figure S2**), and is quite common in both European (non-Finnish MAF 32.5%) and African/African American (MAF 45.9%) populations^54^. The *GCKR* locus was significantly associated with glycated albumin alone (*p* = 4.0 × 10^−8^) as had been previously reported by us^18^. There was no effect size heterogeneity at the *GCKR* lead SNP, yet this locus is nearly significant in European ancestry but not in African ancestry (*p* = 0.74). It is likely due to the limited African ancestry sample size exacerbated by population-level allele frequency differences (non-Finnish European MAF 40.9%, African/African American MAF 13.3%)^54^. The lead SNP at the *FCGRT* locus was absent in our African ancestry genotyped/imputed data (removed due to poor quality) despite common allele frequencies in both European and African populations (non-Finnish European MAF 8.3%, African/African American MAF 30%)^54^. The *FCGRT* locus, implicated in GWAS of serum albumin and lipids^53^, was found to be a novel T2D susceptibility locus, where the lead variant rs142385484– 11 bp away from our lead SNP rs59774409– was genome-wide significant only in a multi-ancestry meta-analysis and not in any single genetic ancestry group.^56^ Although recently implicated as the likely effector transcript for fructosamine through eQTL colocalization of previously-identified locus *RCN3*,^57^ this is the first time *FCGRT* is directly discovered in a GWAS of these biomarkers. The lead variants at *UGT1A1* and *FCGRT* in European ancestry (i.e., 2 of the 5 discovered glycemic trait loci) have moderate to strong evidence of association with T2D (*p* < 10^−3^) in European ancestry-specific meta-analysis and in multi-ancestry meta-analysis^56^ respectively, suggesting that they may be important in T2D pathophysiology (**Table S3**).

### Locus-specific effects at common variants may vary by sex

The sex-specific p-values of multivariate association seemed to indicate that signal at the *ARAP1/STARD10* locus was primarily driven by females while the other loci did not show sex-differentiation (**Table 2**). However, we did not find any significant difference in effect sizes of lead SNPs between females and males at these loci (**Figure S5**). The overall correlation between the sex-stratified p-values at all SNPs in all identified loci was weak regardless of whether BMI was adjusted or not (**Figure S6**). When scanning genome-wide, we did identify locus 13q34 (*TEX29*) was significantly associated in males (rs79276590-C, *p* = 3.0 × 10^−8^) but not in females (**Figure S7**), and exhibited effect size heterogeneity in both biomarkers between sexes (**Figure S8**). This male-specific locus was mapped to gene *TUBGCP3* via 3D chromatin interactions in liver; and no relevant *cis*-eQTL or *cis*-pQTL was found (**Figure S9**). However, the effective sample sizes for genome-wide significant SNPs at this locus were low for males (*n* = 730 to 751) due to really low MAFs and low imputation quality (IMPUTE info scores) in White participants. A few other loci were suggestively significant (5 × 10^−8^ < *p* < 10^−6^) in one sex group but not the other (**Table S4**). In particular, 8q24.11 (*SLC30A8*, a well-known T2D gene^58^) and 19q13.41 (*SIGLEC14*) showed significant sex-differentiation in females, but differed in their effect size heterogeneity (**Figure S10**). We exercise caution in interpreting suggestively significant sex-differentiated loci due to much reduced sample sizes in each group.

**Table 2.** Effects of sex and BMI on the most significant SNPs of the loci identified from multivariate analysis of fructosamine and glycated albumin in multi-ancestry sample. These loci were identified by metaUSAT from sex-combined genotyped/imputed data (either ancestry-stratified or multi-ancestry) at the genome-wide threshold of 5 × 10^−8^.

| Locus | Nearest gene | rsID<br>(lead SNP) | Position<br>(hg19) | Multivariate analysis (metaUSAT) |  |  |  |  |  |
| --- | --- | --- | --- | --- | --- | --- | --- | --- | --- |
|  |  |  |  | BMI-unadjusted p-value |  |  | BMI-adjusted p-value |  |  |
|  |  |  |  | Sex-stratified |  | Sex-combined | Sex-stratified |  | Sex-combined |
|  |  |  |  | Females | Males |  | Females | Males |  |
| 2q37.1 | *UGT1A1* * | rs4148325 | 234673309 | $1.0 \times 10^{-2}$ | $7.5 \times 10^{-5}$ | $5.9 \times 10^{-7}$ | $9.7 \times 10^{-3}$ | $8.1 \times 10^{-5}$ | $2.6 \times 10^{-7}$ |
| 11q13.4 | *ARAPI1*/<br>*STARD10* | rs116714277 | 72473447 | $7.4 \times 10^{-8}$ | $3.4 \times 10^{-2}$ | $2.8 \times 10^{-8}$ | $1.7 \times 10^{-7}$ | $3.2 \times 10^{-2}$ | $3.1 \times 10^{-8}$ |
| 11q22.1 | *RP11-115E19.1* | rs2438321 | 98500410 | $3.2 \times 10^{-1}$ | $9.6 \times 10^{-2}$ | $3.4 \times 10^{-2}$ | $2.4 \times 10^{-1}$ | $1.2 \times 10^{-1}$ | $2.8 \times 10^{-2}$ |
| 17q24.2 | *PRKCA* | rs59443763 | 64526988 | $3.1 \times 10^{-4}$ | $3.5 \times 10^{-5}$ | $1.4 \times 10^{-8}$ | $2.3 \times 10^{-4}$ | $2.3 \times 10^{-4}$ | $6.7 \times 10^{-8}$ |
| 19q13.33 | *FCGRT* | rs59774409 | 50016748 | $1.3 \times 10^{-4}$ | $6.1 \times 10^{-4}$ | $1.3 \times 10^{-7}$ | $3.4 \times 10^{-4}$ | $8.4 \times 10^{-4}$ | $3.0 \times 10^{-7}$ |

### Potential functional genes identified from plasma *cis*-pQTL mapping of detected loci

The lead SNP rs887829 in the *UGT1A1* locus was *cis*-pQTL for proteins encoded by both *UGT1A1* (*p* = 2.0 × 10^−278^, Uniprot ID P22309) and *UGT1A6* (*p* = 5.6 × 10^−104^, Uniprot ID P19224) corresponding to isoforms of the UDP-glucuronosyltransferase 1A protein complex (**Figure 1**). It was identified as *cis*-eQTL for *UGT1A3* and *UGT1A8* in the same complex of alternatively spliced genes. This locus was also mapped to gene *USP40* in T2D-relevant tissues via both eQTL and 3D chromatin mapping strategies in FUMA (the protein UBP40 encoded by *USP40* was not captured in the ARIC pQTL database). The lead SNP rs59774409 of the *FCGRT* locus was *cis*-pQTL for protein encoded by *IRF3* (*p* = 3.4 × 10^−28^, Uniprot ID Q14653) and also *cis*-eQTL for the same gene in adipose tissue (**Figure S11**). No lead SNP identified in African ancestry was a significant *cis*-pQTL. Only one locus contained significant *cis*-pQTLs for protein encoded by *PRKCA*, with rs7222627– 47.3 Kb away from the lead SNP rs59443763– being the most significant *cis*-pQTL (*p* = 3.4 × 10^−29^, Uniprot ID P17252, **Figure 2**). The same gene was mapped in liver tissue via 3D chromatin interactions. A summary of these functional gene prioritizations for the genome-wide significant loci using multiple mapping strategies is provided in **Table 3** and **Figures S12**– **S15**.

**Figure 1.**
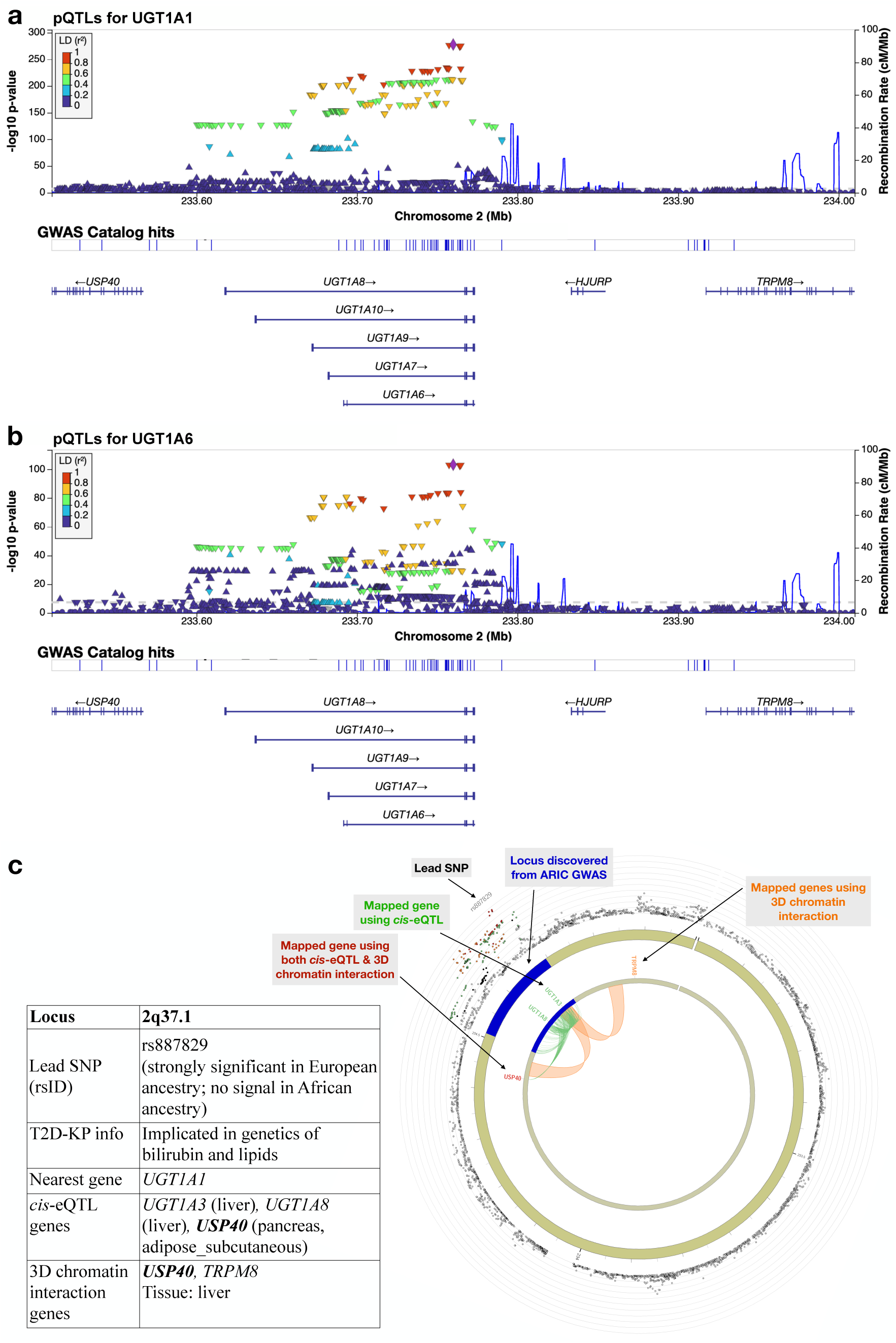
Functional gene prioritization of the chr2q37.1 locus identified from multivariate analysis of fructosamine and glycated albumin using BMI-unadjusted model on sex-combined genotyped/imputed data on European ancestry participants. First two panels show LocusZoom plots of ±250 Kb radius around the lead SNP rs887829 for *cis*-pQTL associations with plasma proteins encoded by (a) *UGT1A1* and (b) *UGT1A6* genes in ARIC White participants from a previous study. (c) This panel summarizes findings from *cis*-eQTL and 3D chromatin interaction mapping strategies in T2D-relevant tissues from external data implemented in FUMA.

**Figure 2.**
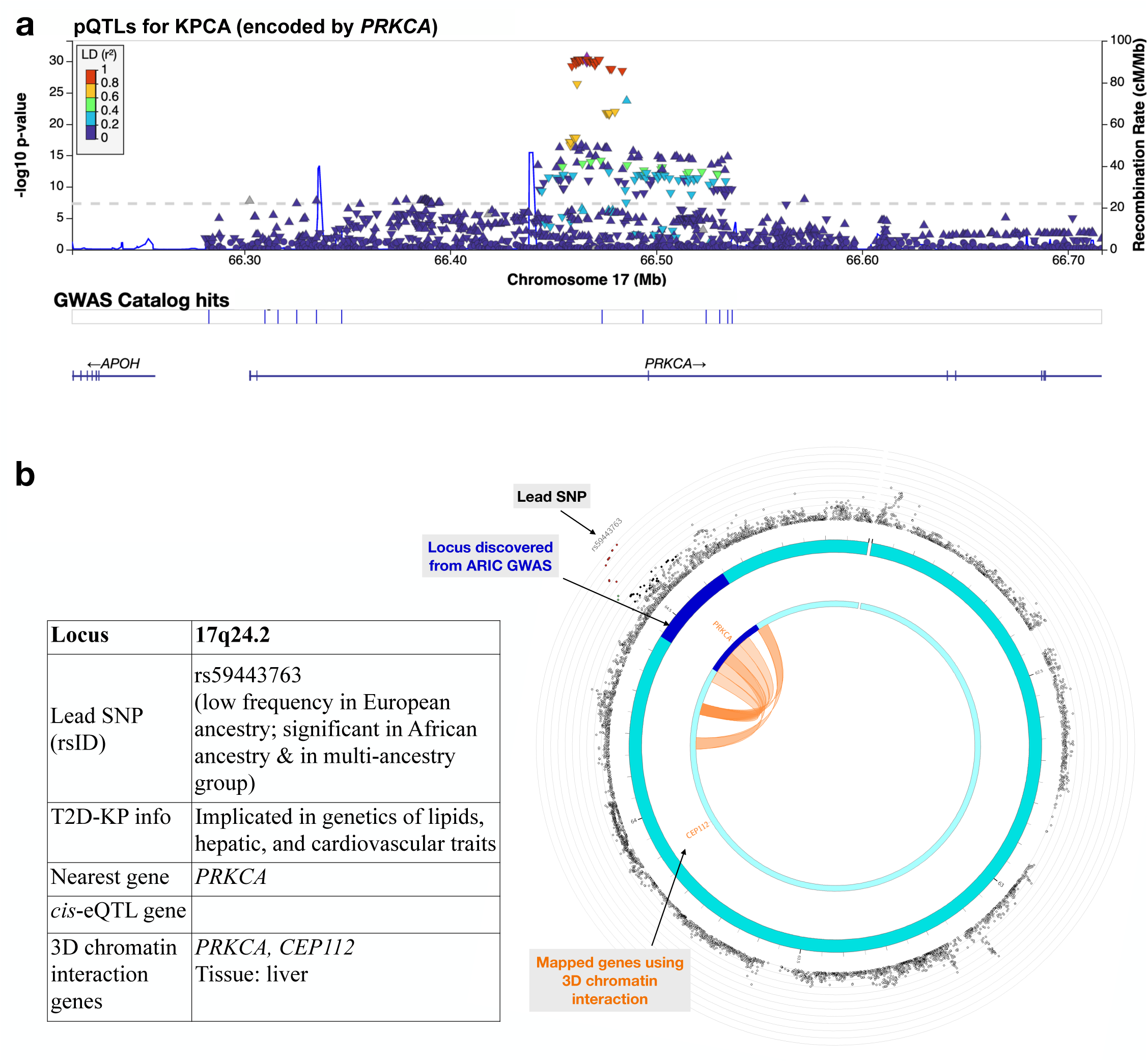
Functional gene prioritization of the chr17q24.2 locus identified from multivariate analysis of fructosamine and glycated albumin using BMI-unadjusted model on sex-combined genotyped/imputed data on African ancestry participants. (a) LocusZoom plot of ±250 Kb radius around the lead SNP rs59443763 for *cis*-pQTL associations with plasma protein encoded by *PRKCA* gene in ARIC Black participants from a previous study. (b) This panel summarizes findings from *cis*-eQTL and 3D chromatin interaction mapping strategies in T2D-relevant tissues from external data implemented in FUMA.

**Table 3.** Functional gene prioritization of the genome-wide significant loci identified from multivariate analysis of fructosamine and glycated albumin using BMI-unadjusted model on genotyped/imputed data. These loci were identified by metaUSAT at the genome-wide threshold of 5 × 10^−8^. The rsID and position corresponds to the most significant SNP (lead SNP) in a locus. The *cis*-pQTL gene mapping used race-stratified ARIC data on plasma proteome from a previously published study. The *cis*-eQTL gene mapping used GTEx v8 data in T2D-relevant tissues and another data source on pancreatic islets. The 3D chromatin interaction gene mapping used Hi-C data in T2D-relevant tissues from GSE87112. Blank cells indicate no significant gene mapping found. See methods section for algorithm we used to classify each locus as potentially “glycemic” or not.

| Locus | Nearest gene | rsID (lead SNP) | Position (hg19) | cis-pQTL gene mapping (plasma) |  | cis-eQTL gene mapping |  | 3D chromatin gene mapping |  | Classified as potentially glycemic? |
| --- | --- | --- | --- | --- | --- | --- | --- | --- | --- | --- |
|  |  |  |  | Gene | Race/ethnicity<sup>†</sup> | Gene | T2D-relevant tissue | Gene | T2D-relevant tissue |  |
| Multi-ancestry (n = 9411) |  |  |  |  |  |  |  |  |  |  |
| 11q13.4<sup>‡</sup> | ARAPI/<br>STARD10 | rs116714277 | 72473447 | – | – | STARD10 | Muscle_skeletal | FCHSD2 | Liver, Pancreas | Glycemic |
|  |  |  |  |  |  |  |  | P2RY2 | Liver |  |
| 17q24.2 | PRKCA | rs59443763 | 64526988 | PRKCA | Black | – | – | PRKCA | Liver | Glycemic |
| European ancestry (n = 7359) |  |  |  |  |  |  |  |  |  |  |
| 2q37.1<sup>‡</sup> | UGT1A1<sup>*</sup> | rs887829 | 234668570 | UGT1A1,<br>UGT1A6 | White | UGT1A3,<br>UGT1A8 | Liver | USP40,<br>TRPM8 | Liver | No |
|  |  |  |  |  |  | USP40 | Adipose_subcutaneous, Pancreas |  |  |  |
| 19q13.33<sup>#</sup> | FCGRT | rs59774409 | 50016748 | IRF3 | White | RPS11 | Adipose_subcutaneous, Adipose_visceral, Muscle_skeletal | – | – | Glycemic |
|  |  |  |  |  |  | FCGRT | Adipose_subcutaneous, Liver, Adipose_visceral |  |  |  |
|  |  |  |  |  |  | RPL13A | Muscle_skeletal, Pancreas |  |  |  |
| African ancestry (n = 2004) |  |  |  |  |  |  |  |  |  |  |
| 11q13.4<sup>‡</sup> | ARAPI/<br>STARD10 | rs116714277 | 72473447 | – | – | STARD10 | Muscle_skeletal | FCHSD2 | Liver, Pancreas | Glycemic |
|  |  |  |  |  |  |  |  | P2RY2 | Liver |  |
| 11q22.1 | RP11-115E19.1 | rs2438321 | 98500410 | – | – | – | – | – | – | Glycemic |
| 17q24.2 | PRKCA | rs59443763 | 64526988 | PRKCA | Black | – | – | PRKCA, CEP112 | Liver | Glycemic |

<sup>†</sup>The *cis*-pQTL results, taken from a previously-published study, were available stratified by self-reported race/ethnicity and not by genetic ancestry.
<sup>‡</sup>Novel locus for fructosamine and/or glycated albumin (may or may not be novel for type 2 diabetes).
<sup>\*</sup>This region includes multiple alternatively spliced genes including *UGT1A1/UGT1A3/UGT1A4/UGT1A5/UGT1A10*.
<sup>#</sup>This locus reported 9 genes in eQTL analysis: *RPS11, FCGRT, RPL13A, PRR12, CPT1C, SCAF1, PRRG2, ALDH16A1, IRF3*; only the top 3 genes with the highest number of eQTLs from this locus are reported in this table.

### Gene-based tests of multivariate phenotype using whole-exome sequence data identify potential effector genes by leveraging rare variants enriched in African ancestry

Across 4 different gene masks (i.e., gene-level variant aggregation approaches) and 2 different kernels– projection and linear kernels for summarizing multivariate phenotype information–, we identified a total of 11 significant genes from gene burden tests performed using GAMuT (**Figure S16**). In particular, when considering common and rare PTVs (Mask 1), the genes *UGDH*, *TXNDC5*, *ARHGEF39*, *C15orf40* and *ZNF208* were significantly associated using one or both phenotype kernels (**Table 4**). Of note, the gene *QSER1* (4 variants, MAC 4, *p*_,-./0123.4_ = 2.3 × 10^−7^, *p*_53406-_ = 3.8 × 10^−8^)– recently implicated in T2D by large-scale studies^45, 59^– was exome-wide significant in our study but failed the minimum MAC gene filter. None of these genes were detected in our stratified analysis due to reduced effective sample size in each ancestry group. We additionally detected *FOSL2* and the region with overlapping genes *RNF103* and *CHMP3* when we added rare missense variants to the set of all PTVs (Mask 2). No gene in this mask was significantly detected in African ancestry; only *RNF103-CHMP3* was significant in European ancestry. By considering pLOFs across the entire allele-frequency spectrum (Mask 3), we identified 4 more significant genes: *CD1D*, *EGFL7*, *MIR126*, *AGPAT2*. We found notable enrichment of pLOFs in African ancestry compared to European ancestry despite the 3-fold smaller sample size for African ancestry in our study. All but *CD1D* also exhibited significant gene-level associations in African ancestry but none in European ancestry. We noted identical findings as Mask 3 when we restricted ourselves to rare pLOFs only (Mask 4). Overall, the QQ plots suggest there could be minor inflation in our gene-based tests for all but Mask 2 (**Figure S17**).

**Table 4.** Gene-based association results from gene burden multivariate analysis of fructosamine and glycated albumin using BMI-unadjusted model on sex-combined, multi-ancestry whole-exome sequence data. These genes were identified by GAMuT from the multi-ancestry data at the exome-wide significance threshold of 2.5 × 10^−6^ either from the projection kernel or the linear kernel or both (here kernels summarize the multivariate phenotype information). Genes with missing p-values in ancestry-stratified analysis are those that fail gene filters (minimum 3 variants and minimum MAC 5).

| Chr | Gene<sup>§</sup> | Multi-ancestry (*n* = 8898) |  |  |  | European ancestry (*n* = 5986) |  |  |  | African ancestry (*n* = 2003) |  |  |  | Classified<br>as<br>potentially<br>glycemic? |
| --- | --- | --- | --- | --- | --- | --- | --- | --- | --- | --- | --- | --- | --- | --- |
|  |  | No. of<br>variants | Total<br>MAC | GAMuT p-value |  | No. of<br>variant<br>s | Total<br>MAC | GAMuT p-value |  | No. of<br>variants | Total<br>MAC | GAMuT p-value |  |  |
|  |  |  |  | Projection<br>kernel | Linear<br>kernel |  |  | Projection<br>kernel | Linear<br>kernel |  |  | Projection<br>kernel | Linear<br>kernel |  |
| Mask 1: protein-truncating variants at any allele frequency |  |  |  |  |  |  |  |  |  |  |  |  |  |  |
| 4 | UGDH | 3 | 5 | 1.2 × 10<sup>−7</sup> | 1.2 × 10<sup>−4</sup> | 3 | 5 | 4.6 × 10<sup>−2</sup> | 2.7 × 10<sup>−2</sup> | 0 | 0 | – | – | No |
| 6 | TXNDC5 | 8 | 5 | 2.2 × 10<sup>−13</sup> | 2.7 × 10<sup>−14</sup> | 3 | 1 | – | – | 5 | 4 | – | – | Glycemic |
| 9 | ARHGEF39 | 9 | 6 | 1.3 × 10<sup>−6</sup> | 2.4 × 10<sup>−7</sup> | 9 | 6 | 1.4 × 10<sup>−1</sup> | 2.3 × 10<sup>−2</sup> | 1 | 1 | – | – | Glycemic |
| 15 | C15orf40 | 3 | 5 | 7.0 × 10<sup>−7</sup> | 5.3 × 10<sup>−3</sup> | 2 | 5 | 5.1 × 10<sup>−2</sup> | 1.6 × 10<sup>−1</sup> | 1 | 1 | – | – | No |
| 19 | ZNF208 | 6 | 7 | 8.1 × 10<sup>−10</sup> | 1.2 × 10<sup>−9</sup> | 5 | 6 | 6.5 × 10<sup>−1</sup> | 8.3 × 10<sup>−1</sup> | 1 | 3 | – | – | Glycemic |
| Mask 2: protein-truncating variants at any allele frequency + missense variants with MAF <5% |  |  |  |  |  |  |  |  |  |  |  |  |  |  |
| 2 | FOSL2 | 34 | 60 | 2.5 × 10<sup>−6</sup> | 1.0 × 10<sup>−6</sup> | 23 | 28 | 2.0 × 10<sup>−1</sup> | 2.4 × 10<sup>−1</sup> | 12 | 39 | 4.7 × 10<sup>−4</sup> | 1.1 × 10<sup>−4</sup> | Glycemic |
| 2 | RNF103-CHMP3 | 23 | 36 | 5.2 × 10<sup>−7</sup> | 9.3 × 10<sup>−5</sup> | 13 | 30 | 1.8 × 10<sup>−7</sup> | 4.5 × 10<sup>−6</sup> | 14 | 13 | 2.4 × 10<sup>−1</sup> | 5.5 × 10<sup>−1</sup> | Maybe |
| Mask 3: putative loss-of-function variants at any allele frequency |  |  |  |  |  |  |  |  |  |  |  |  |  |  |
| 1 | CD1D | 5 | 9 | 1.6 × 10<sup>−7</sup> | 3.3 × 10<sup>−8</sup> | 2 | 2 | – | – | 4 | 7 | 8.9 × 10<sup>−3</sup> | 3.9 × 10<sup>−3</sup> | Glycemic |
| 9 | EGFL7 | 8 | 7 | 8.5 × 10<sup>−11</sup> | 1.2 × 10<sup>−3</sup> | 3 | 2 | – | – | 5 | 6 | 1.6 × 10<sup>−6</sup> | 4.5 × 10<sup>−2</sup> | No |
| 9 | MIR126 | 7 | 6 | 1.5 × 10<sup>−11</sup> | 4.3 × 10<sup>−4</sup> | 3 | 2 | – | – | 4 | 5 | 8.4 × 10<sup>−7</sup> | 3.4 × 10<sup>−2</sup> | No |
| 9 | AGPAT2 | 8 | 7 | 8.5 × 10<sup>−11</sup> | 1.2 × 10<sup>−3</sup> | 3 | 2 | – | – | 5 | 6 | 1.6 × 10<sup>−6</sup> | 4.5 × 10<sup>−2</sup> | Maybe |
| Mask 4: putative loss-of-function variants with MAF <5% |  |  |  |  |  |  |  |  |  |  |  |  |  |  |
| 1 | CD1D | 5 | 9 | 1.6 × 10<sup>−7</sup> | 3.3 × 10<sup>−8</sup> | 2 | 2 | – | – | 4 | 7 | 8.9 × 10<sup>−3</sup> | 3.9 × 10<sup>−3</sup> | Glycemic |
| 9 | EGFL7 | 8 | 7 | 8.5 × 10<sup>−11</sup> | 1.2 × 10<sup>−3</sup> | 3 | 2 | – | – | 5 | 6 | 1.6 × 10<sup>−6</sup> | 4.4 × 10<sup>−2</sup> | No |
| 9 | MIR126 | 7 | 6 | 1.5 × 10<sup>−11</sup> | 4.3 × 10<sup>−4</sup> | 3 | 2 | – | – | 4 | 5 | 8.4 × 10<sup>−7</sup> | 3.4 × 10<sup>−2</sup> | No |
| 9 | AGPAT2 | 8 | 7 | 8.5 × 10<sup>−11</sup> | 1.2 × 10<sup>−3</sup> | 3 | 2 | – | – | 5 | 6 | 1.6 × 10<sup>−6</sup> | 4.5 × 10<sup>−2</sup> | Maybe |

<sup>§</sup>All reported genes are novel for fructosamine and/or glycated albumin (may or may not be novel for type 2 diabetes)

### Classification of detected loci and genes based on their likely biological pathways

Genetic basis of fructosamine and glycated albumin could be influenced through non-glycemic pathways. We classified the significant lead SNPs from our common variant analysis and significant genes from our set-based analyses as potentially “glycemic”, “maybe glycemic” or “non-glycemic” based on considerable effect size attenuation in mediation analysis in ARIC, association analysis with fasting glucose and HbA1c in ARIC, and using single variant and gene-based multi-ancestry meta-analysis results from CMD-KP^50^ (see **Supplementary Methods**). Among the significant loci identified from single variant multi-phenotype analysis of fructosamine and glycated albumin, our algorithm classified *ARAP1/STARD10*, *RP11-115E19.1* and *PRKCA* loci as glycemic as they are likely mediated by fasting glucose, *FCGRT* locus as glycemic due to suggestive evidence of its association with T2D in recent literature, and *UGT1A1* locus as non-glycemic (**Table S5**). From the set-based analyses, the significant genes *TXNDC5*, *ARHGEF39*, *ZNF208*, *FOSL2* and *CD1D* were classified as glycemic while *RNF103-CHMP3* and *AGPAT2* may be glycemic since they were associated with no other glycemic trait but HbA1c (**Table S6**).

## DISCUSSION

This multi-ancestry multi-phenotype analyses using common and rare variants in ARIC study revealed novel genetic underpinnings of the non-traditional glycemic biomarkers fructosamine and glycated albumin. The multi-phenotype analysis of common variants in European ancestry identified the *UGT1A* region and the *FCGRT* locus, which were missed by the typical single-phenotype analysis of fructosamine or glycated albumin alone and by the multi-ancestry joint analysis. The *UGT1A* lead SNP exhibited considerable effect size heterogeneity for fructosamine between European and African ancestries while the *FCGRT* lead SNP was absent in the African ancestry data. The *UGT1A* region is a complex of alternatively spliced genes including *UGT1A1, 1A3, 1A4, 1A5, 1A6, 1A7, 1A8, 1A9* and *1A10* (RefSeq: https://www.ncbi.nlm.nih.gov/nuccore/NM_000463.2), some of which were implicated in our *cis*-pQTL, *cis*-eQTL and 3D chromatin gene mapping strategies. These genes are involved in the glucuronidation of bilirubin (a product of heme catabolism), which creates water-soluble bilirubin. *UGT1A* variants cause the hereditary unconjugated hyperbilirubinemias Crigler-Najjar syndromes and Gilbert syndrome.^60^ In addition, variants in *UGT1A* region are associated with bilirubin levels among individuals without these.^61–63^ Moderately elevated bilirubin is associated with a decreased risk of diabetes and cardiovascular disease.^64–66^ In addition, bilirubin can also bind to albumin.^67^ In the single-phenotype analysis, the variants in *UGT1A* region showed suggestive significance with fructosamine, which is a concentration that does not account for total serum protein. In contrast, variants in *UGT1A* region were not associated with glycated albumin, which is expressed as the percent, accounting for total serum albumin. Because the multivariate phenotype results were likely driven by the fructosamine association, and fructosamine levels are more affected by serum albumin levels, it may be that the multi-phenotype association is impacted by an albumin-related pathway rather than a diabetes-related pathway. If variants in *UGT1A* region affect bilirubin homeostasis, this could alter the amount of albumin bound to bilirubin, which would then impact the amount of albumin available to be glycated, thus impacting fructosamine levels. Nonetheless, we found this locus was genome-wide significant in a multi-ancestry meta-analysis of T2D^56^, suggesting it may be important in T2D pathophysiology.

The joint modeling of fructosamine and glycated albumin using the genotyped/imputed African ancestry data showed three genome-wide significant loci that were also identified in the single-phenotype analyses, and all were likely mediated by fasting glucose. While the *PRKCA* and *RP11-115E19.1* loci have been previously implicated for glycated albumin and fructosamine respectively^18^, the locus in *ARAP1/STARD10* is novel for these glycemic biomarkers. Only the *RP11-115E19.1* locus exhibited significant effect size heterogeneity between ancestries; the lead SNPs for the other two were absent in our European ancestry data due to removal of poor quality SNPs. In the novel locus *ARAP1/STARD10*, also detected in our multi-ancestry analysis, we found significant *cis*-eQTLs for *STARD10* in skeletal muscle, and significant 3D chromatin interactions with *FCHSD2* in liver and pancreas and with *P2RY2* in liver. Common variants in both *ARAP1* and *STARD10* have known associations with T2D and traditional glycemic traits^68–71^. This gene-rich region encompassing *ARAP1*, *STARD10*, and *FCHSD2* is strongly associated with T2D in CMD-KP^50^, which along with our mediation analysis indicates this locus likely influences these non-traditional glycemic traits via a diabetes-related pathway.

Multi-ancestry joint analysis using variant-sets from the exome sequencing data revealed 11 new genes associated with these non-traditional glycemic biomarkers. In particular, 5 genes– *TXNDC5*, *ARHGEF39*, *ZNF208*, *RNF103-CHMP3* and *CD1D*– were also significantly associated with fasting glucose or HbA1c in ARIC. *UGDH* is involved in starch and sucrose metabolism, pentose and glucuronate interconversions; the Human Protein Atlas (HPA)^72^ indicates its tissue specificity (liver) and its involvement in lipid metabolism in intestine and liver based on gene expression clustering; and there is ‘moderate genetic support’ for involvement in 2-hour insulin^50^ based on Human Genetic Evidence (HuGE) score^73^. *TXNDC5* plays an important role in iron metabolism (CMD-KP shows ‘very strong genetic support’ for hemoglobin concentration), and appears to play a role in diabetes progression and response of pancreatic cells to high glucose exposure^74^. There is ‘extreme genetic support’ for involvement of *ZNF208* and ‘moderate support’ for *FOSL2*, particularly due to rare variants, in fasting insulin adjusted for BMI and 2-hour insulin respectively.^50^ There is some evidence in literature showing role of *FOSL2* in insulin regulation and glucose metabolism in humans and mice.^75^ CMD-KP shows ‘very strong genetic support’ for *RNF103* in BMI, and the HPA indicates its involvement in metabolism in liver based on gene expression clustering.

Rare pLOFs have the potential to elucidate gene function^40^, and we found 4 such significant genes. *CD1D* could influence our non-traditional glycemic biomarkers via both diabetes- and blood-related pathways as evidenced from associations of rare pLOFs in *CD1D* with fasting glucose (*p* = 2.2 × 10^−6^) and HbA1c (*p* = 1.8 × 10^−5^) in ARIC, ‘very strong’ HuGE score in favor of both HbA1c and red blood cell distribution width in CMD-KP, and multiple common variants in close proximity to this gene were strongly associated with blood traits in Open Targets Genetics portal^76, 77^. A ‘moderate genetic support’ for involvement of rare variants in *EGFL7* in glycemic trait adiponectin and of common variants in *EGFL7* in BMI-adjusted fasting insulin are indicative of this gene’s likely influence via a glycemic pathway.^50^ *EGFL7* may also act via a non-glycemic pathway, particularly a blood-related pathway, as implicated by ‘very strong’ HuGE score for involvement in lymphocyte count^50^ and common variant associations with blood traits^77^. There is some evidence of association of dysregulation of microRNA-126 (miR-126, the RNA type for *MIR126* gene) with T2D and related complications^78–80^ but *MIR126* is not associated with fasting glucose or HbA1c in ARIC and little is known about this gene. *AGPAT2* may be a glycemic gene (in CMD-KP, HbA1c rare variant gene-based analysis *p* = 0.003) but its role in T2D is unclear.

A major limitation of this work is lack of validation in an external dataset. Fructosamine and glycated albumin are not routinely collected in epidemiologic studies. Lack of studies with both genome-wide data on diverse ancestry and rigorous measurements of these biomarkers currently precludes a larger GWAS than the current study. However, by leveraging joint patterns, correlations and genetic overlap between biomarkers, and by exploiting genotyped and exome sequence data to query variants across the entire allele frequency spectrum, we were able to improve power to unravel genetic architecture of fructosamine and glycated albumin. We compensated, to some extent, the lack of replication data by assessing the role of discovered loci and genes in T2D using existing literature and large-scale public databases such as CMD-KP. Our approach for classifying discovered loci/genes as potentially “glycemic” based on attenuation of effect estimates when adjusted for fasting glucose is not perfect since fasting glucose measurements are prone to error and is not a great gold standard. Further follow-up of the potentially glycemic loci/genes from this study is necessary to help uncover mechanisms by which these genetic regions may influence risk to T2D. It will be an area of future work to investigate pleiotropy between the traditional and these non-traditional glycemic biomarkers, and investigate causal relationships among these biomarkers by leveraging the loci not involved in a diabetes-related pathway. Nonetheless, this study expands the inventory of loci and genes associated with fructosamine and glycated albumin (some of which are unique to these glycemic biomarkers and others that overlap with T2D-susceptibility regions), finds evidence of potential cross-ancestry differences in biology of these biomarkers (e.g., heterogenous effect sizes at the level of genetic ancestry), and finds suggestive evidence of sex-specific effects.

## Data Availability Statement

The genotyped data and the whole exome sequence data from ARIC are available in dbGaP (https://www.ncbi.nlm.nih.gov/projects/gap/cgi-bin/study.cgi?study_id=phs000090.v7.p1 and https://www.ncbi.nlm.nih.gov/projects/gap/cgi-bin/study.cgi?study_id=phs000668.v5.p1 respectively).

## Data Availability

The genotyped data and the whole exome sequence data from ARIC used in this study are available in dbGaP.

https://www.ncbi.nlm.nih.gov/projects/gap/cgi-bin/study.cgi?study_id=phs000090.v7.p1

## Acknowledgments

All analyses were carried out using computing cluster—the Joint High Performance Computing Exchange—at the Department of Biostatistics, Johns Hopkins Bloomberg School of Public Health. We thank Dr. Candelaria Vergara at the Johns Hopkins Bloomberg School of Public Health for her guidance with the identification of genetic ancestry of study participants. The authors are thankful to the staff and the participants of the ARIC Study for their important contributions.

## Author Contributions

Conceptualization: DR, SJL, ES, PD

Data curation: SV, AT

Formal analysis: DR, SJL

Funding acquisition: DR, ES, PD

Investigation: DR, SJL

Methodology: DR, SJL

Project administration: DR, SJL

Software: DR

Supervision: DR, NC, ES, PD

Validation: DR

Visualization: DR

Writing – original draft: DR, SJL

Writing – review & editing: DR, SJL, SV, AT, BY, NC, ES, PD

## Web Resources

ARIC pQTL summary statistics, http://nilanjanchatterjeelab.org/pwas/

GAMuT, http://www.biostat.umn.edu/∼baolin/research/gamut/

Human Protein Atlas, https://www.proteinatlas.org/

metaUSAT (v1.17), https://github.com/RayDebashree/metaUSAT

PLINK (v1.9), https://www.cog-genomics.org/plink/

SNP2GENE (v1.3.7), FUMA https://fuma.ctglab.nl/

SnpEff (v5.0e), https://pcingola.github.io/SnpEff

