## Supplementary for "Characterizing common and rare variations in non-traditional glycemic biomarkers using multivariate approaches on multi-ancestry ARIC study"

### SUPPLEMENTARY METHODS

#### Genotyping, imputation, and quality control

Samples were genotyped on the Affymetrix 6.0 array and imputed to 1000 Genomes Phase I (March 2012) separately by race/ethnicity using IMPUTE2. Samples failing quality control from mismatches in self-reported and genetic sex, genetic outliers, failed concordance with previous TaqMan genotypes, first-degree relatedness with another study member and missingness $> 98\%$ were excluded. SNPs with missingness $>$ 5%, or Hardy-Weinberg Equilibrium (HWE) $p < 10^{-5}$ were excluded. Further, SNPs that had duplicated basepair positions, minor allele frequency (MAF) <5%, IMPUTE info score <0.8 representing poor imputation quality, and indels were removed. Hard calls (A,T,G,C) were obtained from imputed dosages for analysis. The final dataset included 6,792,306 SNPs in the sample of White participants and 5,313,666 SNPs in the sample of Black participants.

#### Exome sequencing and quality control

Whole exome sequencing was done as part of the Cohorts for Heart and Aging Research in Genomic Epidemiology (CHARGE) consortium at the Baylor College of Medicine Human Genome Sequencing Center (HGSC) using DNA extracted from blood. Samples were sequenced on the Illumina HiSeq 2000 or 2500 platform (San Diego, CA), and quality control measures implemented. Single nucleotide variants (SNVs) were excluded if they met any of the following criteria: posterior probability $< 0.95$, variant read count $< 3$, variant read ratio $< 0.25$ or $> 0.75$, strand bias $> 99\%$ in single direction, total convergence $< 10 \times$ for SNVs ($< 30 \times$ for indels), outside exon capture regions, monomorphic variant, missing rate $>$ 20%, mappability score $< 0.8$, mean depth coverage $> 500 \times$, HWE $p < 5 \times 10^{-6}$ in ancestry-specific groups. Samples were excluded if they had $> 20\%$ missing data or beyond 6 standard deviations from the mean read depth, singleton count, heterozygote to homozygote ratio, or transition to transversion (Ti/Tv) ratio. First-degree related individuals were removed. The post quality control sample included 7,810 White participants and 3,180 Black participants with genetic data on 2,556,859 SNVs and 76,133 indels.

#### Identification of genetic ancestry of ARIC participants

We stratified all ARIC participants either by European or by African genetic ancestry rather than by self-reported race. We determined genetic ancestry based on tight clustering of ARIC participants with 1000 Genomes superpopulations in the top three genetic principal components (PC) space. To do this, we first merged the ARIC genetic array data on 9,747 White and 3,207 Black individuals with 1000 Genomes Phase 3 v5 data on 2,504 individuals (there were 800,382 genetic variants common to both datasets). After removing variants with 3 or more alleles and those with strand mismatch, there were a total of 797,454 variants in common. For generating the genetic PCs, we further removed variants with MAF $< 1\%$, genotyping rate or non-missing genotype data $< 95\%$, and HWE p-value $< 10^{-3}$, possible phase mismatch and then pruned them using LD $R^2 > 0.1$, window of 50 bp with step size 5 bp.

Next, we generated genetic PCs by implementing PC-AiR[1] on the merged dataset with 47,317 filtered and pruned variants. For PC calculation, we excluded 970 related individuals who were identified as having estimated kinship coefficient $> 2^{-7/2}$ within each group by KING-robust estimator[2] (note, unlike PCs for ancestry determination, all analyses in this manuscript were done after removing first-degree related individuals among the ARIC participants with the pertinent biomarker and genetic data). Only individuals most representative of genetic ancestry of the group were retained. PCs were calculated on unrelated set of individuals, and afterwards related individuals were projected onto the PC space (**Figure S0a**). White and

*Figure S0. Distribution of ARIC White and Black participants on the PC space with 1000 Genomes superpopulations, and determination of genetic ancestry of ARIC participants.*

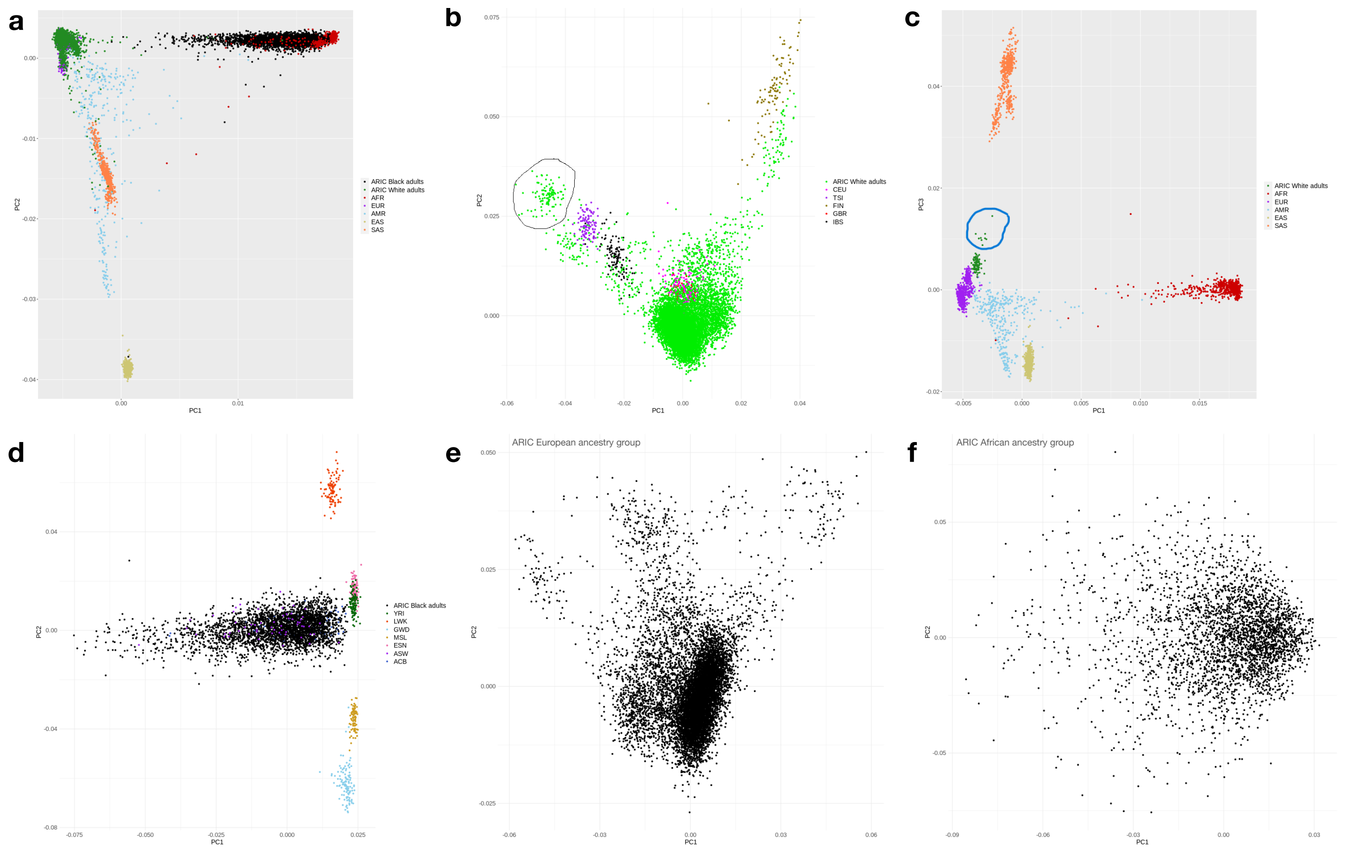


Black ARIC individuals that respectively "clustered closely" with 1000G AFR and EUR superpopulations were selected. We defined "close clustering" by visually inspecting the PC space: White individuals were selected if they fell in the space defined by $PC_1 \leq -0.0025$ and $PC_2 \geq -0.003$ while Black individuals were selected if they satisfied $PC_1 \geq 0.0025$ and $PC_2 \geq 0$.

The selected White ($n = 9{,}700$) and Black ($n = 3{,}187$) individuals were separately mapped onto a new PC space along with 1000G EUR and AFR superpopulations respectively. We found that except one small group, all selected White individuals ($n = 9{,}596$) clustered well with all 1000G EUR sub-populations (**Figure S0b**). We further investigated this small group of White individuals ($n = 104$) not clustering well by looking at not only $PC_1$-$PC_2$ plot of these individuals with all 1000G populations but also $PC_2$-$PC_3$, $PC_1$-$PC_3$ and the 3-dimensional $PC_1$-$PC_2$-$PC_3$ plots. Except a few White individuals ($n = 9$), the rest clustered well with 1000G EUR superpopulation on the $PC_1$-$PC_3$ space (**Figure S0c**). On the other hand, all selected Black individuals ($n = 3{,}187$) clustered well with 1000G ASW (African Ancestry in Southwest US) population while the remaining continental African ancestry groups clustered away (**Figure S0d**).

The above 9,691 White and 3,187 Black individuals that clustered well with EUR and AFR superpopulations were further filtered (we removed obvious outliers and individuals falling 6 standard deviations away from the means of $PC_1$ and $PC_2$ – recommended/default choice in EIGENSOFT package[3,4]) and projected onto new PC spaces separately without any 1000G individuals. We iterated this process of filtering and re-calculating PCs until no ARIC individual can be removed (**Figure S0e-f**). Finally, we classified 9,524 White and 3,170 Black individuals from ARIC as European and African genetic ancestry groups respectively. The Pearson's correlation coefficient between MAFs from this ARIC European ancestry group and MAFs from 1000G EUR individuals was 0.99. Similarly, the Pearson's correlation coefficient between this ARIC African ancestry group and 1000G AFR was 0.97.

In our final dataset, among the 7,395 White ARIC participants with genotyped/imputed data and non-missing data on our glycemic biomarkers of interest (fructosamine and percent glycated albumin), 7,359 were identified as belonging to European ancestry. Similarly, 2,004 of the 2,016 Black ARIC participants were identified as belonging to African ancestry. Among the 6,589 White ARIC participants with whole exome sequence data and non-missing glycemic biomarker data, 5,986 were identified as belonging to European ancestry. Similarly, 2,003 of the 2,309 Black ARIC participants were identified as belonging to African ancestry.

**Statistical association analyses: more details**

***Single variant multi-phenotype analysis.***

The usual approach of analyzing one phenotype at a time ignores the phenotypic correlation structure and their relatedness which is the equivalent to discarding information, and is burdened by multiple testing correction at both the variant and the phenotype level. While there are several methods for multi-phenotype (multivariate) analysis, the optimality of them highly depends on underlying true nature of complex pleiotropic associations[5]. Data-adaptive multivariate methods ensure robust power performance across different alternatives[6], that vary from one variant to the next. One such method is Unified Score-based Association Test (USAT)[5], a data-adaptive test of association of multiple continuous phenotypes with a single genetic variant. We used metaUSAT[7], the version of USAT that uses single-phenotype single variant GWAS summary statistics with improved computational efficiency (R program v1.17, https://github.com/RayDebashree/metaUSAT). For a given genetic variant, metaUSAT gives weights to two types of multi-phenotype methods: metaMANOVA (a multivariate linear regression model implementation using GWAS summary statistics) and a marginal score test (based on single-phenotype regression models). It, then, adaptively selects the weighted test with minimum p-value and computes an approximate asymptotic p-value of multivariate association.[5, 7] In the Results section, any mention of population-level allele frequencies of lead SNPs (most significant SNP in a locus) is taken from gnomAD v2.1.1 browser[8].

***Rare variant set-based multi-phenotype analysis.***

A single variant test of multiple phenotypes is not ideal for exome sequence data due to the large number of low-frequency and rare variants. Multi-phenotype single variant methods (e.g., metaUSAT) can be prone to spurious association signals at low MAFs due to possible violation of multivariate normality assumption for the joint distribution of either biomarkers or the single-phenotype GWAS summary statistics.[6] We therefore implemented gene-based multivariate analysis of the biomarkers on variants with MAF$<$ 5% from the exome sequence data using GAMuT[9], which performs a non-parametric test of no association between a set of phenotypes and a set of genetic variants. It summarizes the set of phenotypes (or a set of genotypes in a gene or a region of interest) and their interdependence using a phenotype (or genotype) similarity matrix. It then assesses if pairwise phenotypic similarity is independent of pairwise genotypic similarity in the sample.

Rare variants predicted to cause loss of functions (pLOFs) and deleterious missense variants have the potential to elucidate gene function.[10] Similar to previous human exome sequencing studies[11-13], we defined 4 different, sometimes nested, sets of functionally important rare and common variants called variant masks using SnpEff[14] (v5.0e, https://pcingola.github.io/SnpEff): (1) protein-truncating variants (PTVs) at any allele frequency with annotations frameshift_variant, initiator_codon_variant, splice_acceptor_variant, splice_donor_variant, stop_lost, stop_gained; (2) PTVs in Mask 1 plus missense variants with MAF<5%; (3) pLOFs at any allele frequency, as annotated by SnpEff and extracted by SnpSift[15] (inclusion criterion: all entries having LOF with genes that have >50% transcripts affected); (4) pLOFs with MAF<5%. We also created a mask involving rare deleterious missense variants (inclusion criterion: all entries having missense annotation and putative impact/deleteriousness 'HIGH') and rare pLOFs; however, the resultant mask was identical to Mask 4 involving only rare pLOFs. Gene regions were defined in SnpEff using GRCh37 Ensembl release 75.

To assess cumulative effect of variants in a gene, we collapsed all variants into a single burden genotype as done previously[10]. Briefly, for a gene as defined by a specific gene mask, (a) individuals who carry zero copies of minor allele for all variants in that gene are assigned genotype value 0; (b) individuals who carry one copy of minor allele for at least one variant in that gene are assigned genotype value 1; (c) the rest are assigned genotype value 2. We used an updated version of GAMuT in MSKAT[16] package (http://www.biostat.umn.edu/~baolin/research/gamut/) for more accurate small p-value computation. For the biomarkers, we considered phenotype similarity matrix using both projection and linear kernels (i.e., two different ways of summarizing multivariate phenotype information). For each mask, we only included genes with at least 3 qualifying variants and a burden minor allele count (MAC) of 5 or more[17]. For our multi-ancestry analysis, Masks 1-4 respectively contained 4882, 17726, 1396, and 1375 genes. We used a common, conservative Bonferroni-corrected exome-wide significance threshold of $2.5 \times 10^{-6}$ for all gene-based tests (4 masks × 2 phenotype similarity matrices).

**Functional gene prioritization**

In the Results section, for ease of referencing, we annotated identified loci with gene names based on proximity to the lead SNPs regardless of evidence of causal gene, if any, in the literature. Recent articles on GWAS SNP to causal gene mapping have implicated that the closest gene is usually the causal gene[10, 18, 19]. As described below, we also considered other gene mapping strategies for functional gene prioritization.

To characterize the regulatory effects of the significant signals and prioritize functional genes, we searched for overlap between detected genetic determinants from our single variant multi-phenotype analysis and plasma protein quantitative trait loci (pQTLs). We used the race-stratified pQTL summary statistics in ARIC from a previously published study[20]. Note, stratification by race (as done in the ARIC pQTL study) is not the same as stratification by genetic ancestry (as done in this study). US individuals identifying as Black are enriched for European and African genetic ancestries, while US individuals identifying as White are enriched for European genetic ancestry.[21]

We additionally used three strategies in FUMA to perform functional gene mapping of the identified loci: positional mapping, eQTL mapping, and 3D chromatin interaction mapping. For positional gene mapping of a locus based on ANNOVAR[22] annotations, we used a maximum distance of 10 Kb between SNPs and genes. For eQTL gene mapping, we used GTEx v8 expression data[23] from T2D-relevant tissues (Adipose_Subcutaneous, Adipose_Visceral, Liver, Muscle_Skeletal, Pancreas)[24], and transcript expression data from human pancreatic islets[25]. Only SNP-gene pairs significant at FDR 5% were used and we did not filter any SNP by functional annotation for eQTL mapping. Note, 85.3% of individuals from GTEx v8 are of European ancestry[20], and thus eQTL findings may be limited for loci identified from African ancestry participants in this study. For chromatin interaction mapping, we used Hi-C data from liver and pancreas tissues in GSE87112[26] and declared significant chromatin interaction at FDR $\leq 10^{-6}$ (default suggested threshold). Genes whose promoter regions (defined as 250bp upstream and 500bp downstream of TSS) overlapped with Hi-C interacted region were used for gene mapping. Note, only four donors contributed tissues for the Hi-C data whose ancestry information are not publicly available.[27]

**Characterization of detected loci and genes as glycemic vs non-glycemic: more details**

***Loci identified from single variant multi-phenotype analysis.***

Genetic basis of fructosamine and glycated albumin could be influenced through non-glycemic pathways. To understand biological pathways of these biomarkers that relate to glycemia, we repeated all our analyses by conditioning on fasting glucose and followed a classification algorithm similar to those used previously[28,29]. Briefly, we classified a locus as "glycemic" if (a) effect size estimates of its lead SNP for both fructosamine and glycated albumin are reduced by at least 25% when fasting glucose is adjusted in the model, thus indicating a glycemic pathway mediated by fasting glucose, or (b) the lead SNP is associated ($p < 10^{-4}$) with any glycemic trait except HbA1c (since many genetic associations of HbA1c are often driven through associations with blood cell traits[28]) in a phenome-wide association (PheWAS) analysis of all traits available in Common Metabolic Diseases Knowledge Portal (CMD-KP)[30] as of July 22, 2022. We considered the following glycemic traits (unadjusted and adjusted for BMI wherever available) from CMD-KP: fasting glucose, 2-hour glucose, random glucose, fasting insulin, 2-hour insulin, insulin at 30-min OGTT, fasting proinsulin, and T2D. Note, the PheWAS results were generated by "bottom-line" integrated analysis across all datasets in CMD-KP, which means sample overlaps between pairs of studies/datasets were estimated and appropriately accounted for before conducting multi-ancestry meta-analysis.[31] The choice of 25% attenuation threshold was previously suggested as achieving optimal balance between sensitivity and specificity.[28]

If association effect sizes were unchanged or increased in the mediation analysis and not associated with any glycemic trait in CMD-KP, we used single-phenotype association results with fasting glucose ($p < 0.005$) in ARIC to classify them as "glycemic". Here we chose a liberal threshold of $p < 0.005$ following existing literature[29] because we have a modest sample size of <10,000 in ARIC while effective sample sizes for CMD-KP were often >500,000. For the remaining unclassified variants, we marked the lead SNP as "maybe glycemic" if associated with HbA1c in CMD-KP ($p < 10^{-4}$) or in ARIC ($p < 0.005$), else marked them as "non-glycemic". Both fasting glucose and HbA1c measurements were from ARIC visit 2 (1990-1992) and we obtained their single-phenotype association results (multi-ancestry as well as ancestry-stratified results) as described in "Statistical association analyses" section before. It is possible for a lead SNP to be associated with both glycemic traits and other non-glycemic traits (e.g., blood-related traits) in CMD-KP; we classified such variants as "glycemic" since in our classification we are primarily concerned about loci affecting fructosamine and glycated albumin in a manner not reflecting ambient glycemia[28].

### SUPPLEMENTARY FIGURES

**Figure S1:** Sample inclusions and exclusions in our analyses.

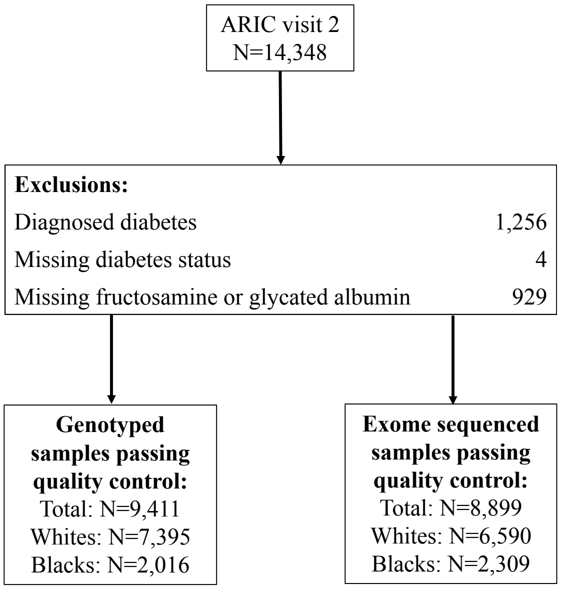

**Figure S2:** Heterogeneity in effect sizes of fructosamine and glycated albumin at the lead SNPs of loci from ancestry-stratified analysis summarized in Table 1. Here effect size refers to mean change in log(biomarker) for every additional copy of the effect allele. The number in parenthesis after genetic ancestry label is the number of non-missing genotypes (sample size) used to obtain the effect size. A missing ancestry group in any plot indicates that the variant is either monomorphic or not present in that group in our sample.

*(a) European ancestry*

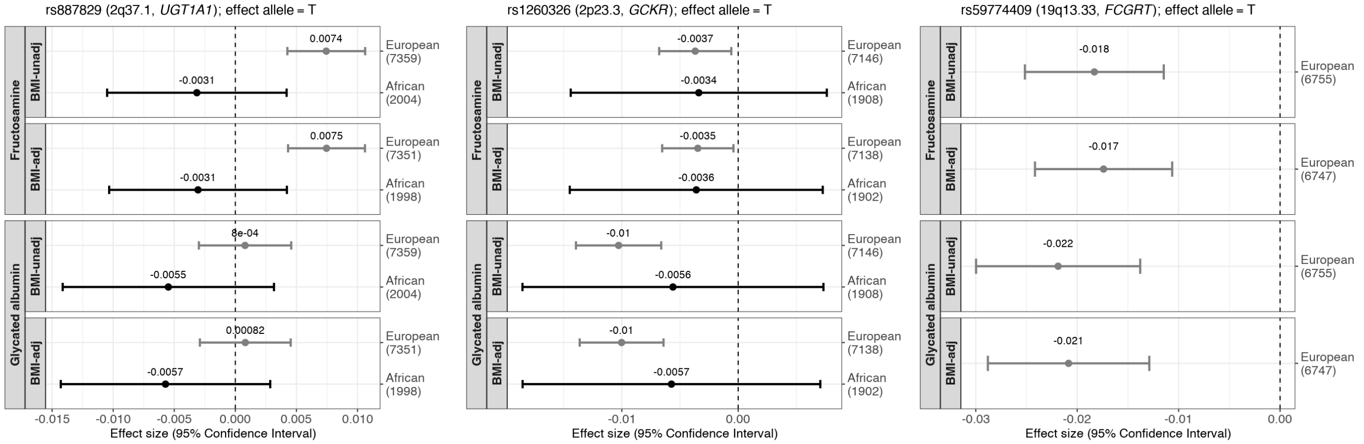


*(b) African ancestry*

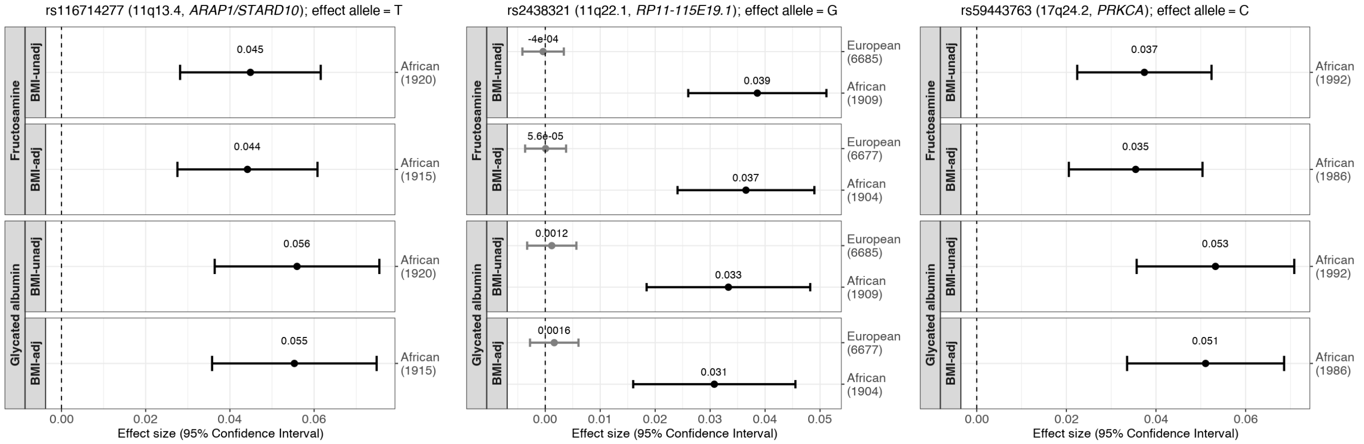

**Figure S3:** Common variant associations from multivariate analysis of fructosamine and glycated albumin using metaUSAT on genotyped/imputed data from sex-combined multi-ancestry data. The red horizontal line corresponds to the genome-wide significance threshold of $p < 5 \times 10^{-8}$. (a) Manhattan plot for the BMI-unadjusted model. (b) Manhattan plot for the BMI-adjusted model. (c) Scatter plot of p-values from BMI-unadjusted and adjusted models for the SNPs that were suggestively significant at $10^{-5}$ in both.

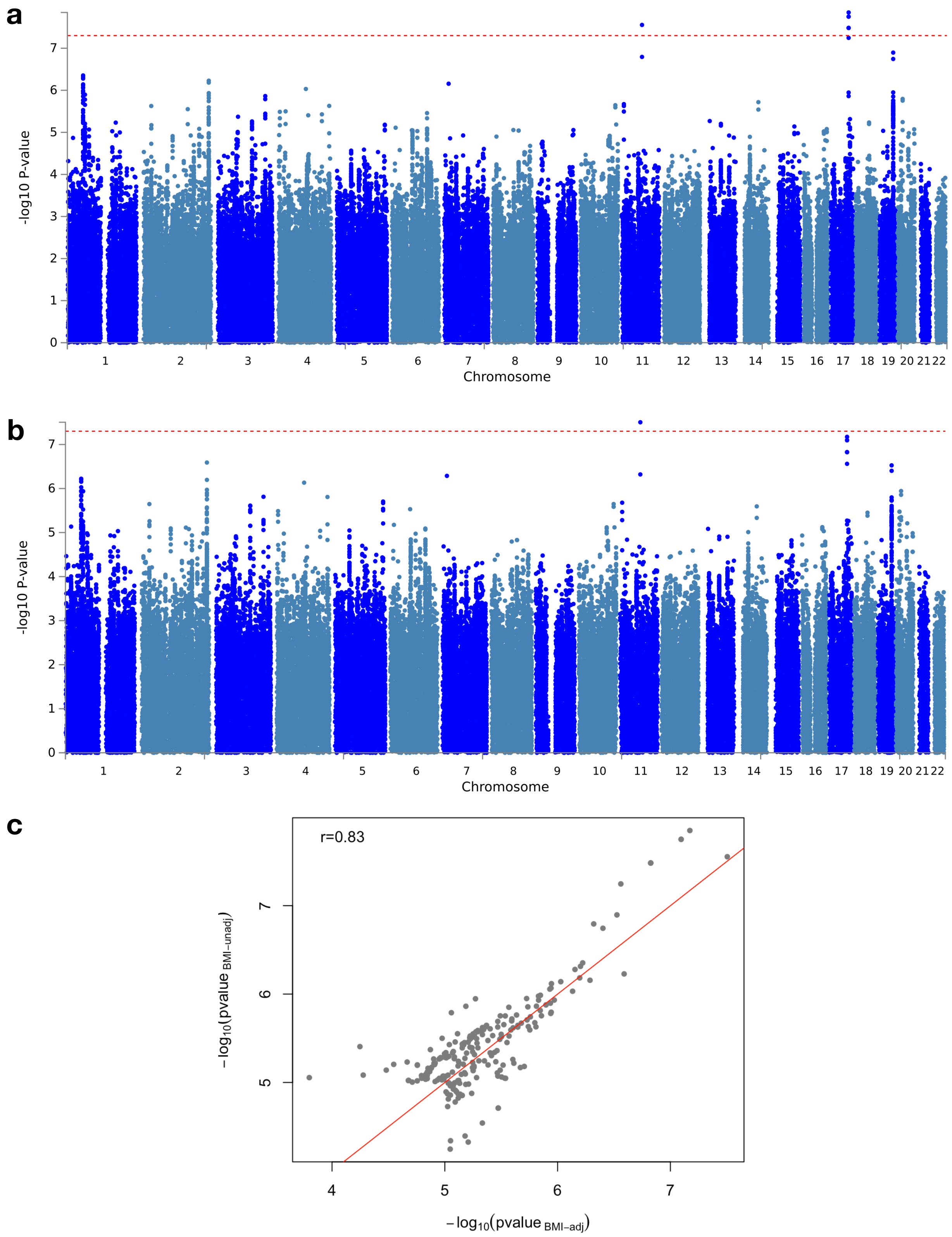

**Figure S4:** Common variant associations from multivariate analysis of fructosamine and glycated albumin using metaUSAT on sex-combined and ancestry-stratified genotyped/imputed data. The red horizontal line corresponds to the genome-wide significance threshold of $p < 5 \times 10^{-8}$.
(a) Manhattan plots for the BMI-unadjusted (left) and adjusted models (right) for European ancestry only.
(b) Manhattan plots for the BMI-unadjusted (left) and adjusted models (right) for African ancestry only.

(a) *European ancestry*

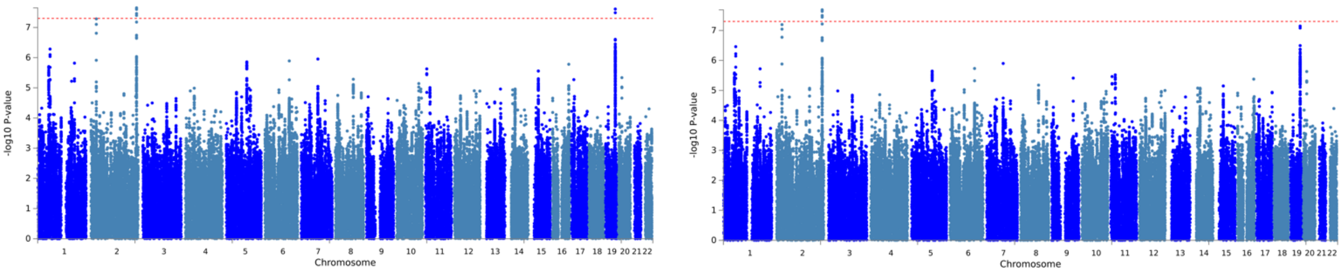


(b) *African ancestry*

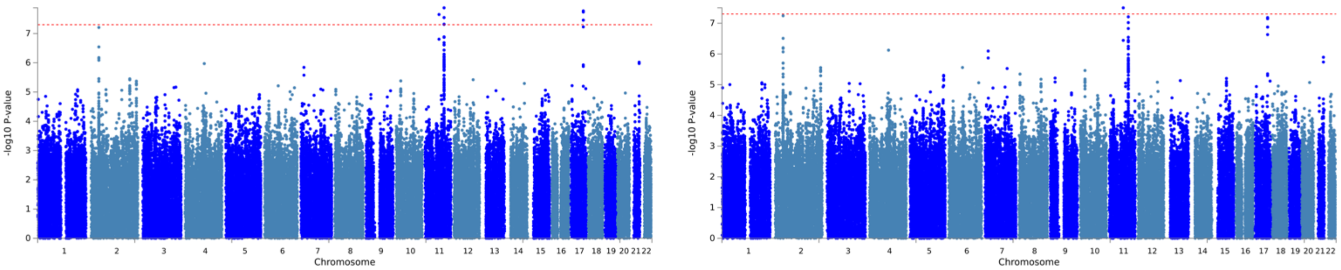

**Figure S5:** Heterogeneity in effect sizes of fructosamine and glycated albumin for females and males at the lead SNPs of loci (summarized in Table 1) based on multi-ancestry data. These loci were identified by metaUSAT from sex-combined multi-ancestry genotyped/imputed data at a suggestive threshold of $10^{-6}$. Here effect size refers to mean change in log(biomarker) for every additional copy of the effect allele. The number in parenthesis after sex is the number of non-missing genotypes (sample size) used to obtain the effect size for a given sex.

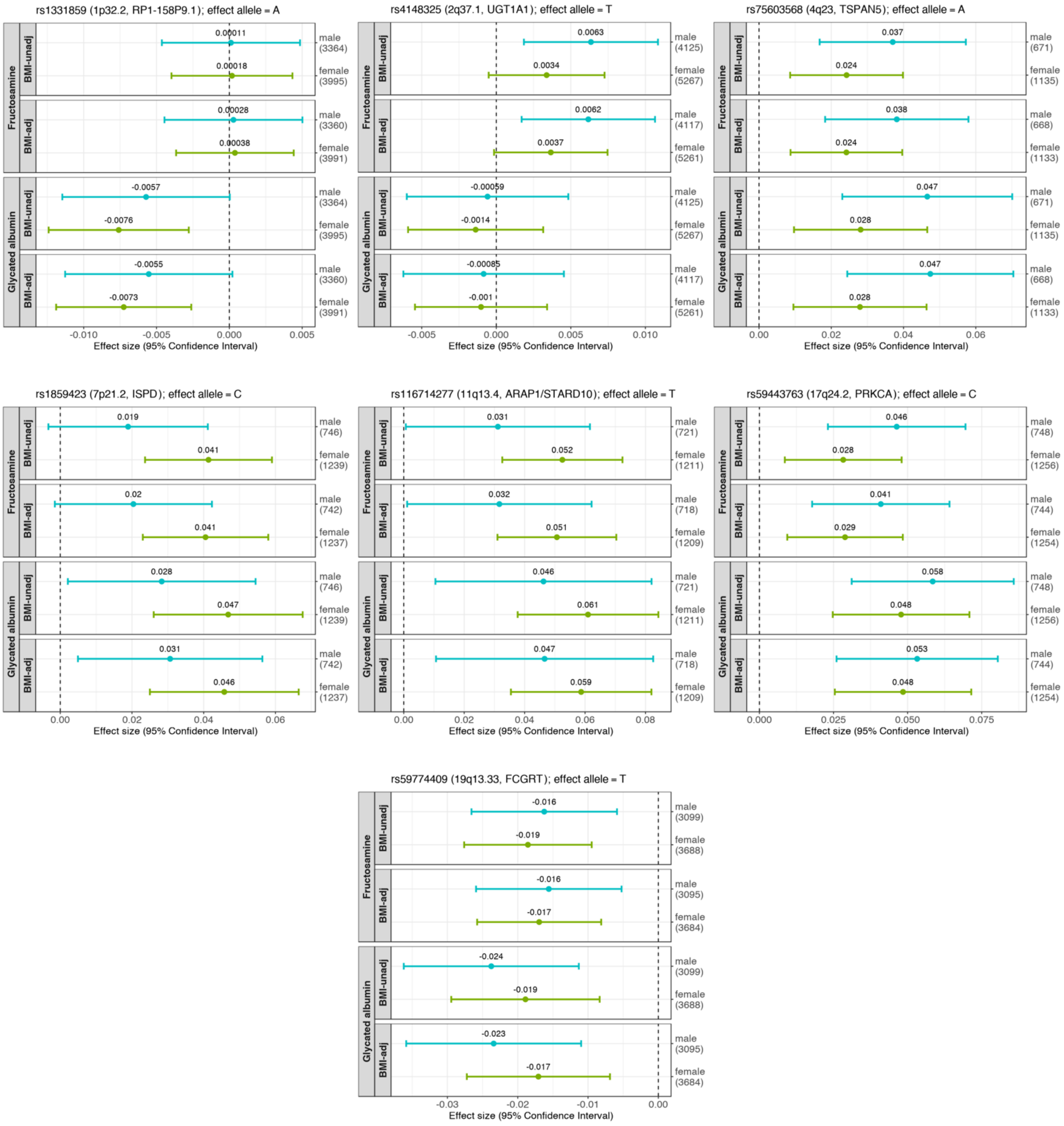

**Figure S6:** Heterogeneity in effects of sex and BMI on the loci identified from multivariate analysis of fructosamine and glycated albumin. These loci were identified by metaUSAT from genotyped/imputed data on sex-combined multi-ancestry data at a suggestive threshold of $10^{-6}$.

*Heterogeneity in effects of sex*

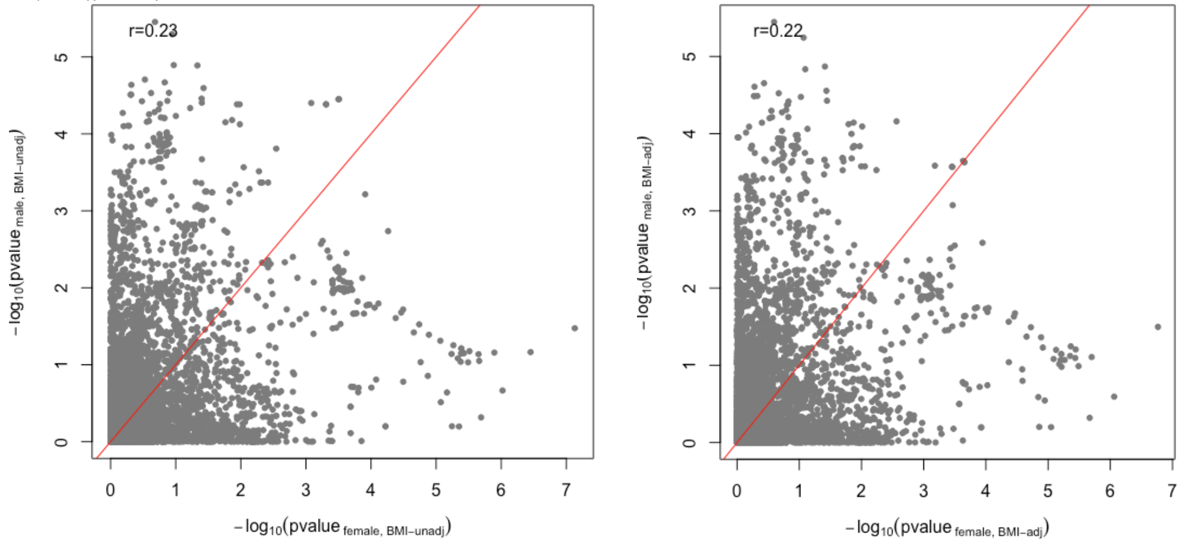


*Heterogeneity in effects of BMI*

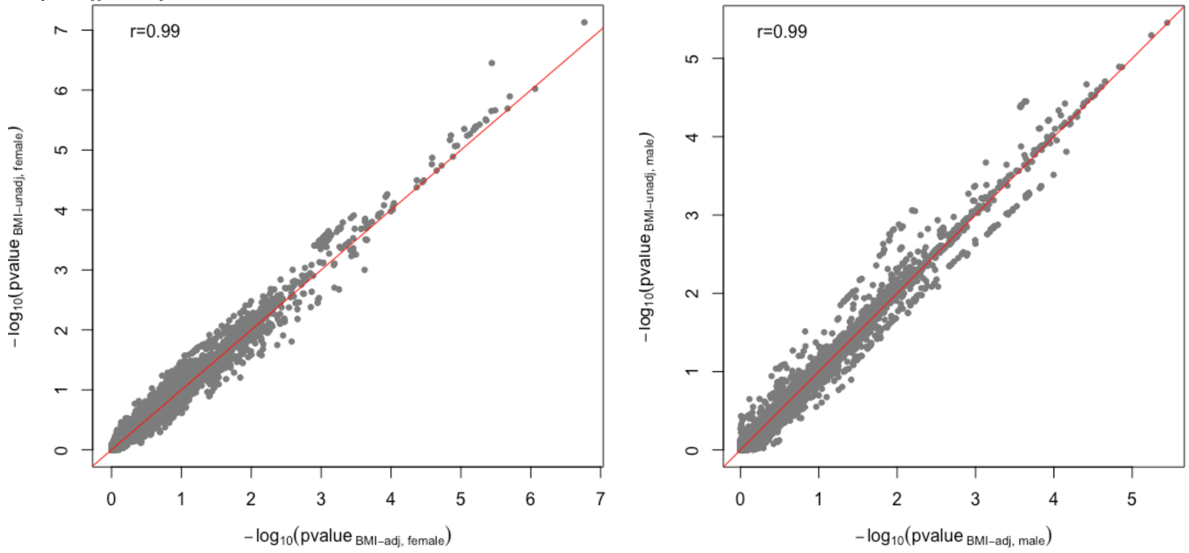

**Figure S7:** Common variant associations from multivariate analysis of fructosamine and glycated albumin using metaUSAT on sex-stratified multi-ancestry genotyped/imputed data. The red horizontal line corresponds to the genome-wide significance threshold of $p < 5 \times 10^{-8}$.
(a) Manhattan plots for the BMI-unadjusted (left) and adjusted (right) models for females only.
(b) Manhattan plots for the BMI-unadjusted (left) and adjusted (right) models for males only.

(a) *Females*

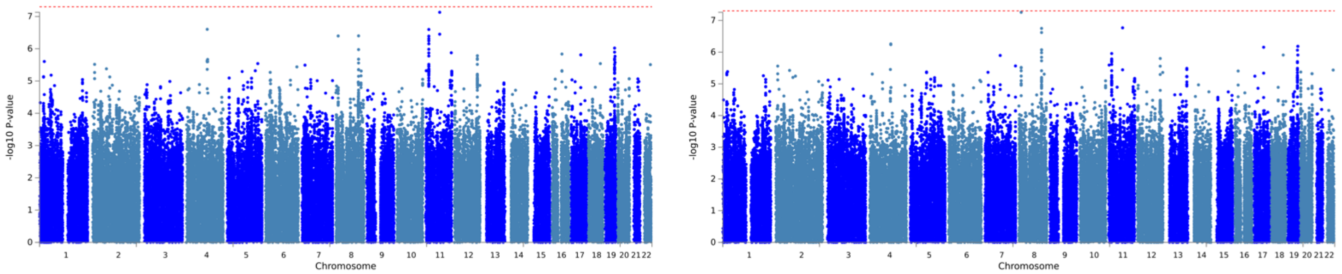


(b) *Males*

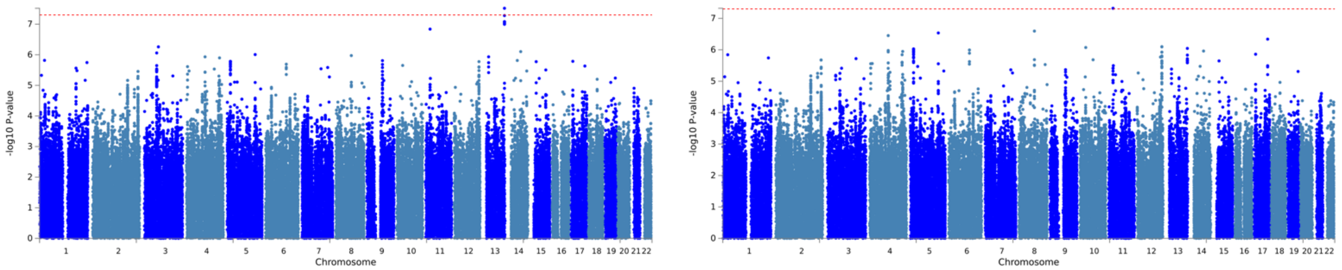

**Figure S8:** Heterogeneity in effect sizes of fructosamine and glycated albumin for females and males at the lead SNPs of **loci identified in males only** (summarized in Table S4). These loci were identified by metaUSAT from multi-ancestry genotyped/imputed data on males at a suggestive threshold of $10^{-6}$. Here effect size refers to mean change in log(biomarker) for every additional copy of the effect allele. The number in parenthesis after sex is the number of non-missing genotypes (sample size) used to estimate the effect size for a given sex.

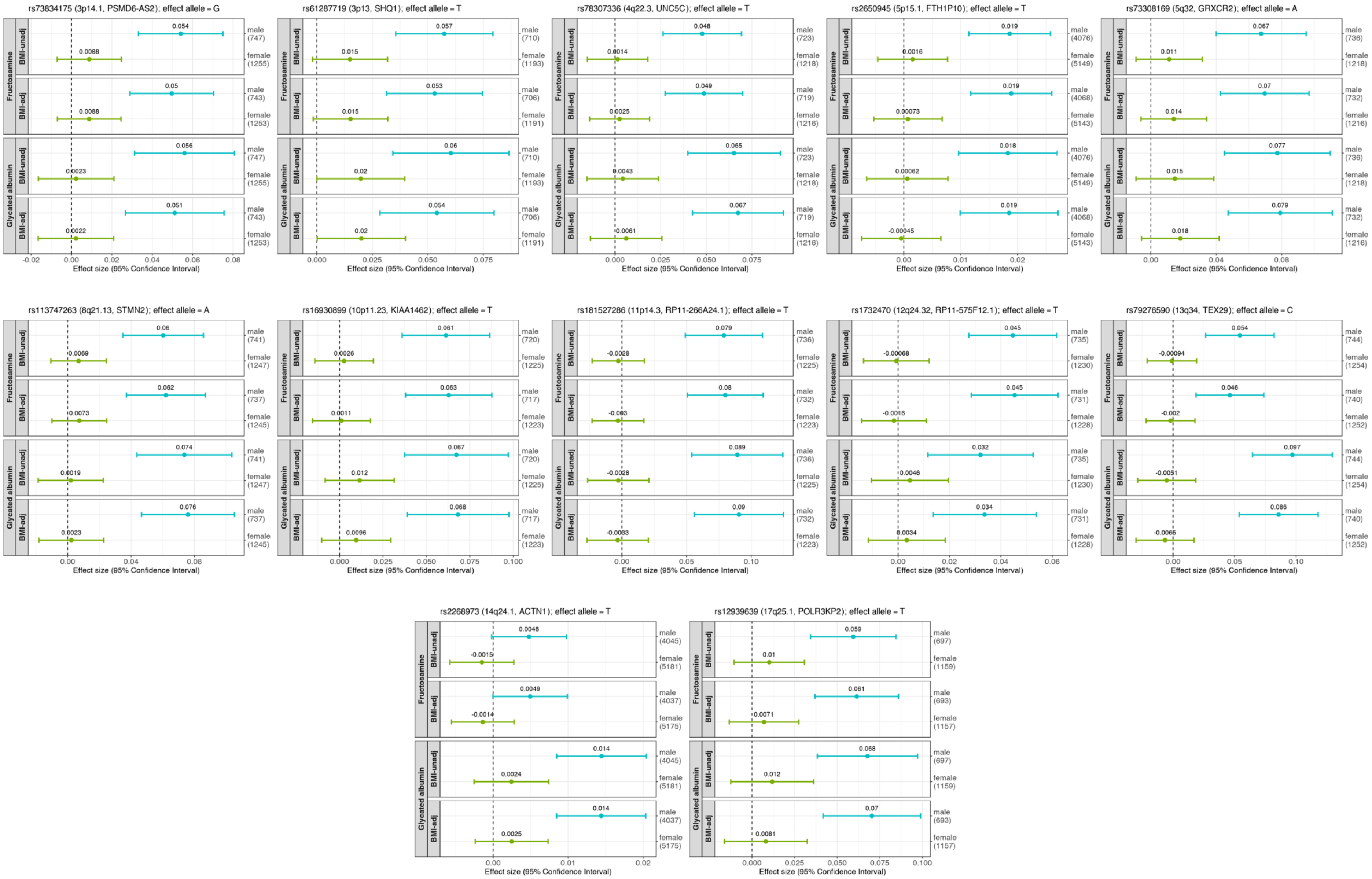

**Figure S9:** Regional plot of the **chr13q34** locus genome-wide significant in multivariate analysis of fructosamine and glycated albumin using BMI-unadjusted model on **multi-ancestry** genotyped/imputed data on **males**. The panels show GWAS association statistics, CADD score, Regulome DB score, eQTL and 3D chromatin interaction information from FUMA. The *cis*-eQTL gene mapping used GTEx v8 data in T2D-relevant tissues and another data source on pancreatic islets. The 3D chromatin interaction gene mapping used Hi-C data in T2D-relevant tissues from GSE87112.

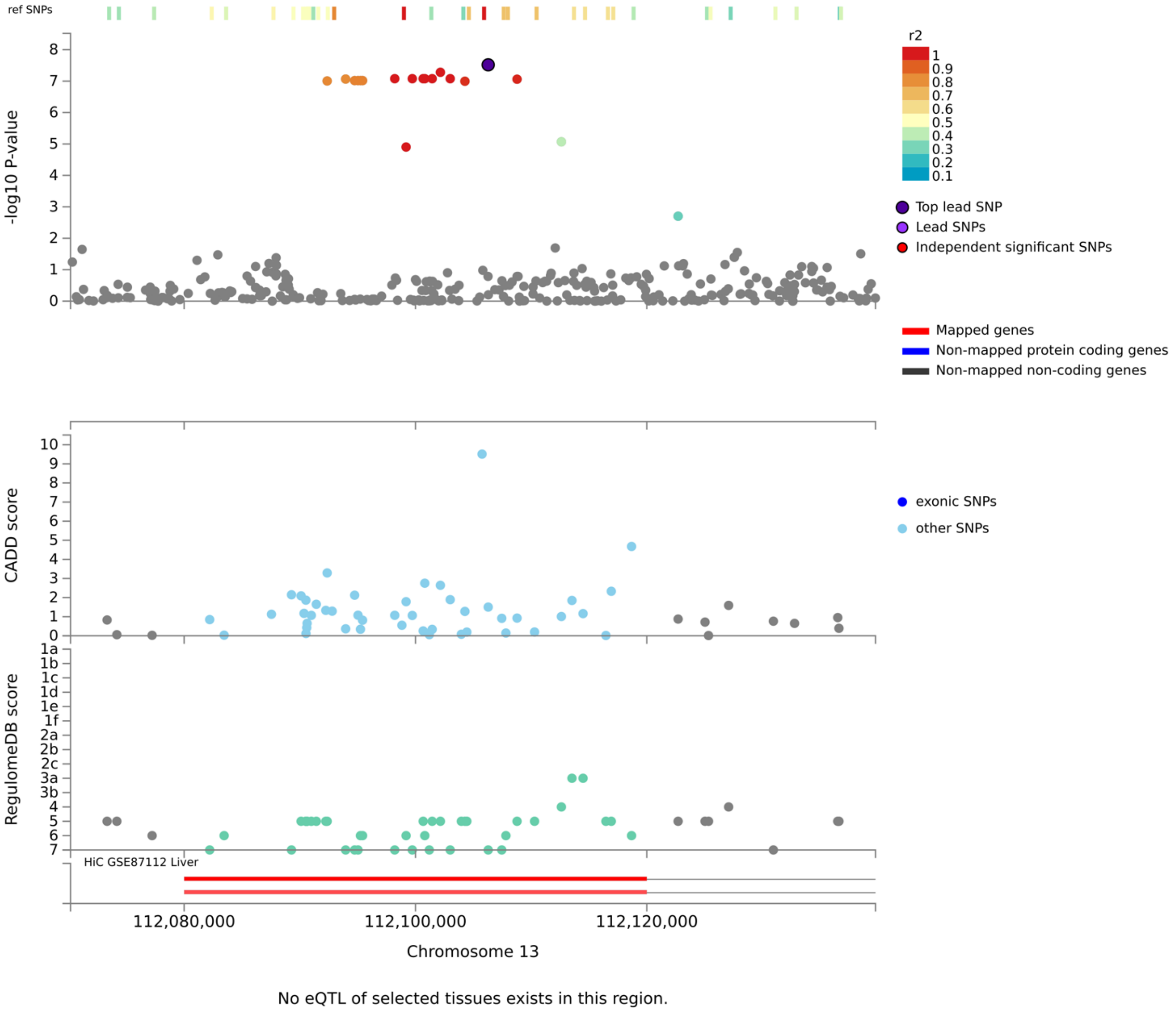


| Locus | Nearest gene | rsID | Position (hg19) | *cis*-pQTL gene mapping (plasma) | | *cis*-eQTL gene mapping | | 3D chromatin gene mapping | |
|---|---|---|---|---|---|---|---|---|---|
| | | | | Gene | Race/ ethnicity | Gene | T2D-relevant tissue | Gene | T2D-relevant tissue |
| 13q34 | *TEX29* | rs79276590 | 112106288 | – | – | – | – | *TUBGCP3* | Liver |

**Figure S10:** Heterogeneity in effect sizes of fructosamine and glycated albumin for females and males at the lead SNPs of **loci identified in females only** (summarized in Table S4). These loci were identified by metaUSAT from multi-ancestry genotyped/imputed data on females at a suggestive threshold of $10^{-6}$. Here effect size refers to mean change in log(biomarker) for every additional copy of the effect allele. The number in parenthesis after sex is the number of non-missing genotypes (sample size) used to estimate the effect size for a given sex.

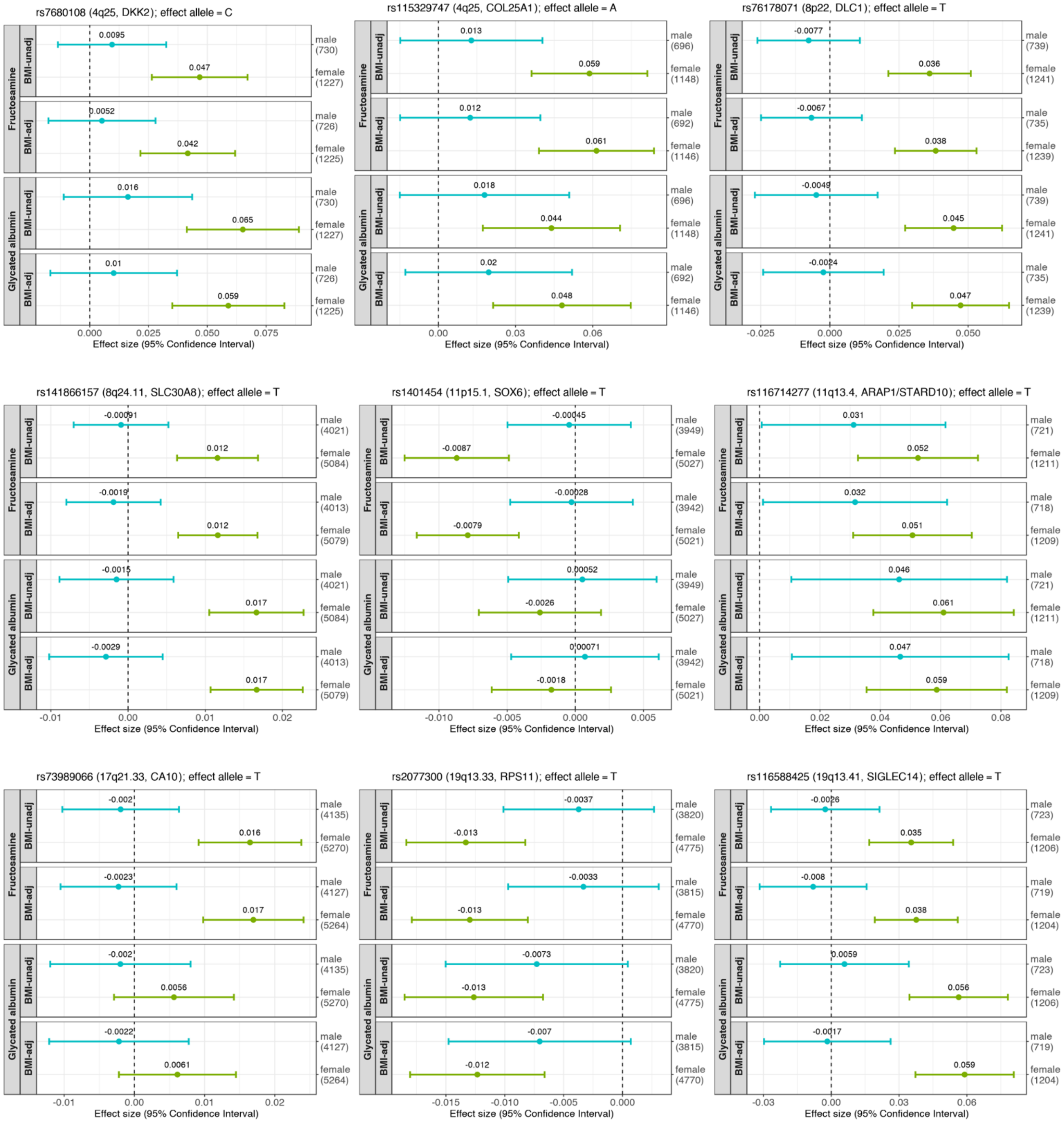

**Figure S11:** Functional gene prioritization of the **chr19q13.33** locus identified from multivariate analysis of fructosamine and glycated albumin using BMI-unadjusted model on **sex-combined**, genotyped/imputed data on **European ancestry** participants. First two panels show LocusZoom plots of ±250 Kb radius around the lead SNP rs59774409 for *cis*-pQTL associations with plasma proteins encoded by (a) *DKKL1* and (b) *IRF3* genes in White ARIC participants from a previously published study. (c) This panel summarizes findings from *cis*-eQTL and 3D chromatin interaction mapping strategies in T2D-relevant tissues from external data implemented in FUMA.

a

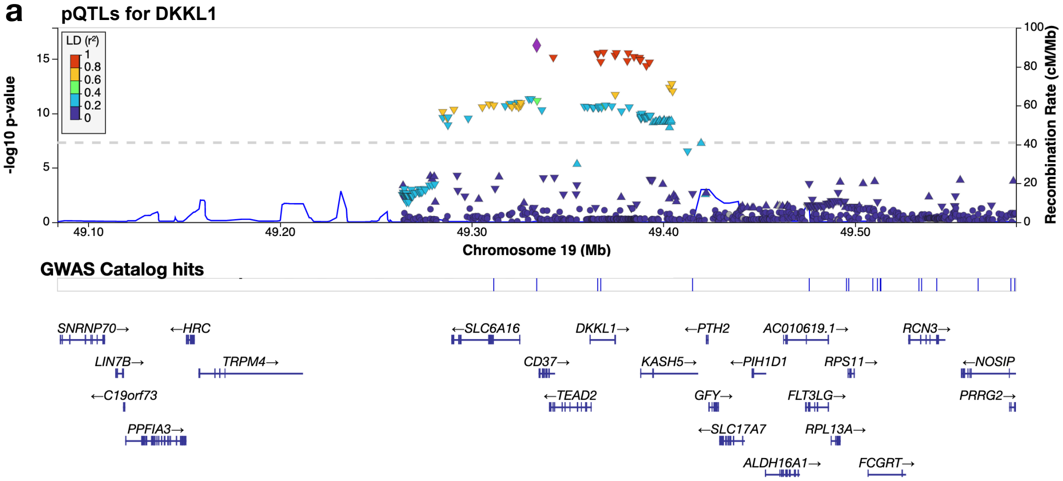


b

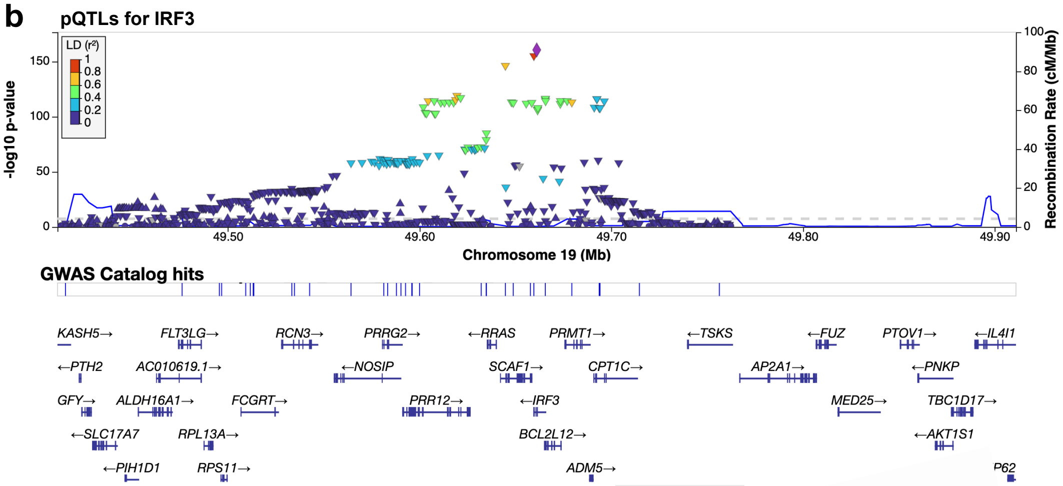


c

| Locus | 19q13.33 |
|---|---|
| Lead SNP (rsID) | rs59774409 (low frequency in African ancestry; suggestively significant in European ancestry & multi-ancestry group) |
| T2D-KP info | Implicated in genetics of serum albumin, calcium metabolite, lipids |
| Nearest gene | *FCGRT* |
| *cis*-eQTL genes | Adipose: *RPS11, FCGRT, CPT1C, SCAF1, PRR12, IRF3*<br>Muscle skeletal: *RPS11, RPL13A, CPT1C, ALDH16A1*<br>Liver: *FCGRT, PRRG2, PRR12*<br>Pancreas: *PRR12, RPL13A, ALDH16A1* |
| 3D chromatin interaction gene | |

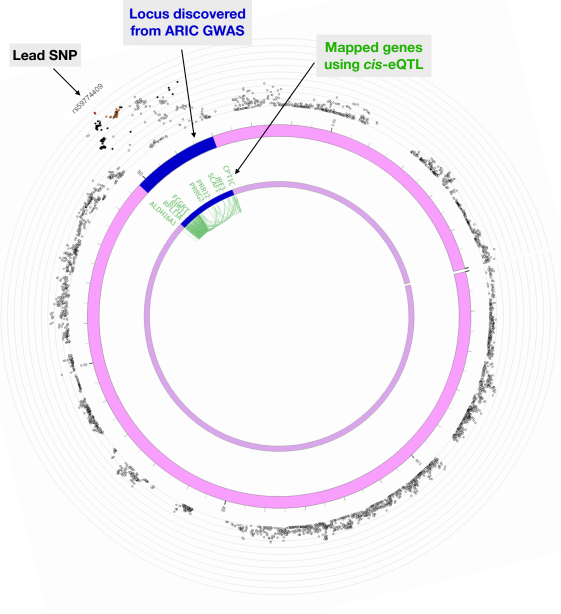

**Figure S12:** Regional plot of the **chr2q37.1** locus genome-wide significant in multivariate analysis of fructosamine and glycated albumin using BMI-unadjusted model on **sex-combined** genotyped/imputed data on **European ancestry** participants. The panels show GWAS association statistics, CADD score, Regulome DB score, eQTL and chromatin interaction information from FUMA. The *cis*-eQTL gene mapping used GTEx v8 data in T2D-relevant tissues and another data source on pancreatic islets. The 3D chromatin interaction gene mapping used Hi-C data in T2D-relevant tissues from GSE87112.

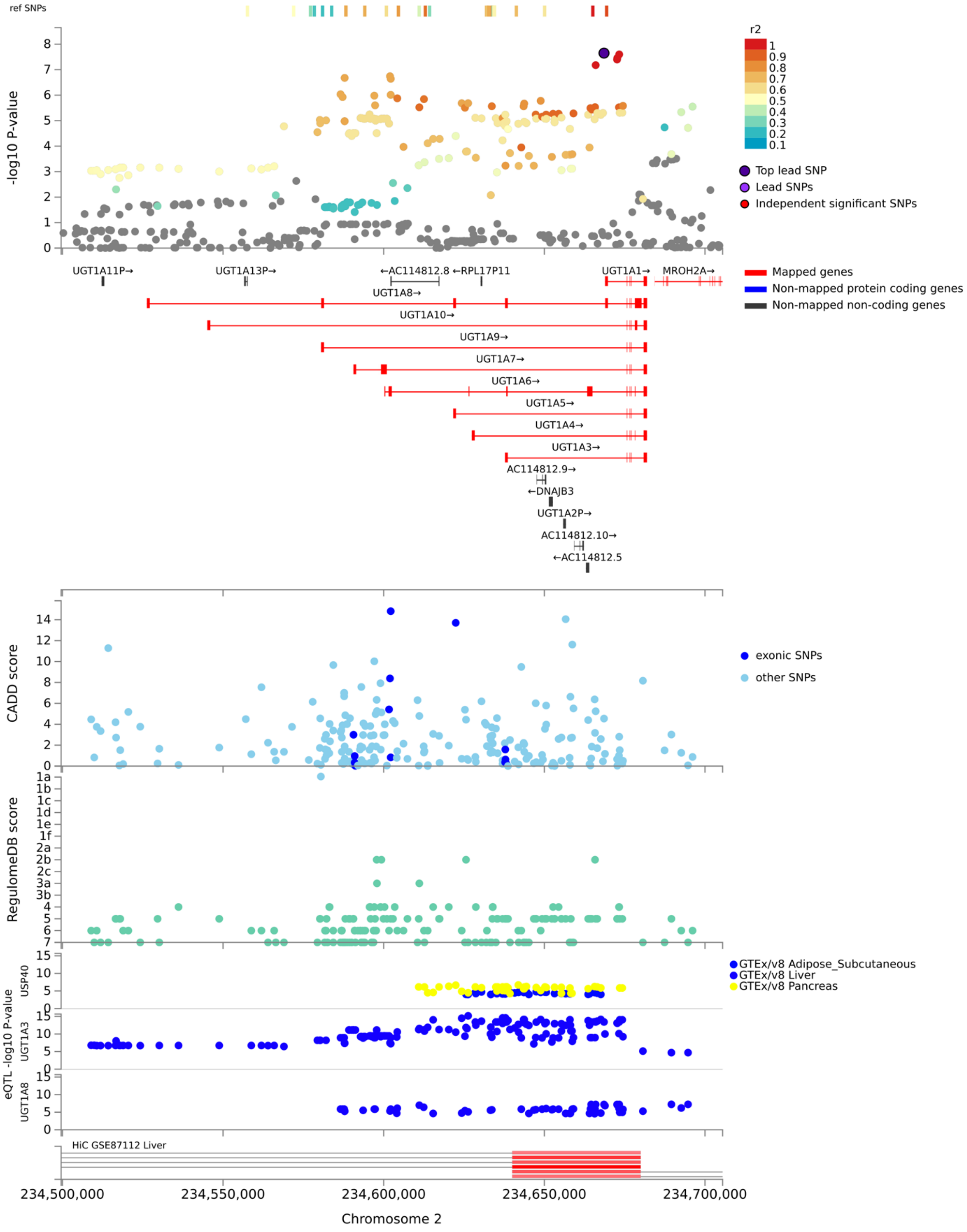

**Figure S13:** Regional plot of the **chr11q13.4** locus genome-wide significant in multivariate analysis of fructosamine and glycated albumin using BMI-unadjusted model on **sex-combined** genotyped/imputed data on **African ancestry** participants. The panels show GWAS association statistics, CADD score, Regulome DB score, eQTL and chromatin interaction information from FUMA. The *cis*-eQTL gene mapping used GTEx v8 data in T2D-relevant tissues and another data source on pancreatic islets. The 3D chromatin interaction gene mapping used Hi-C data in T2D-relevant tissues from GSE87112.

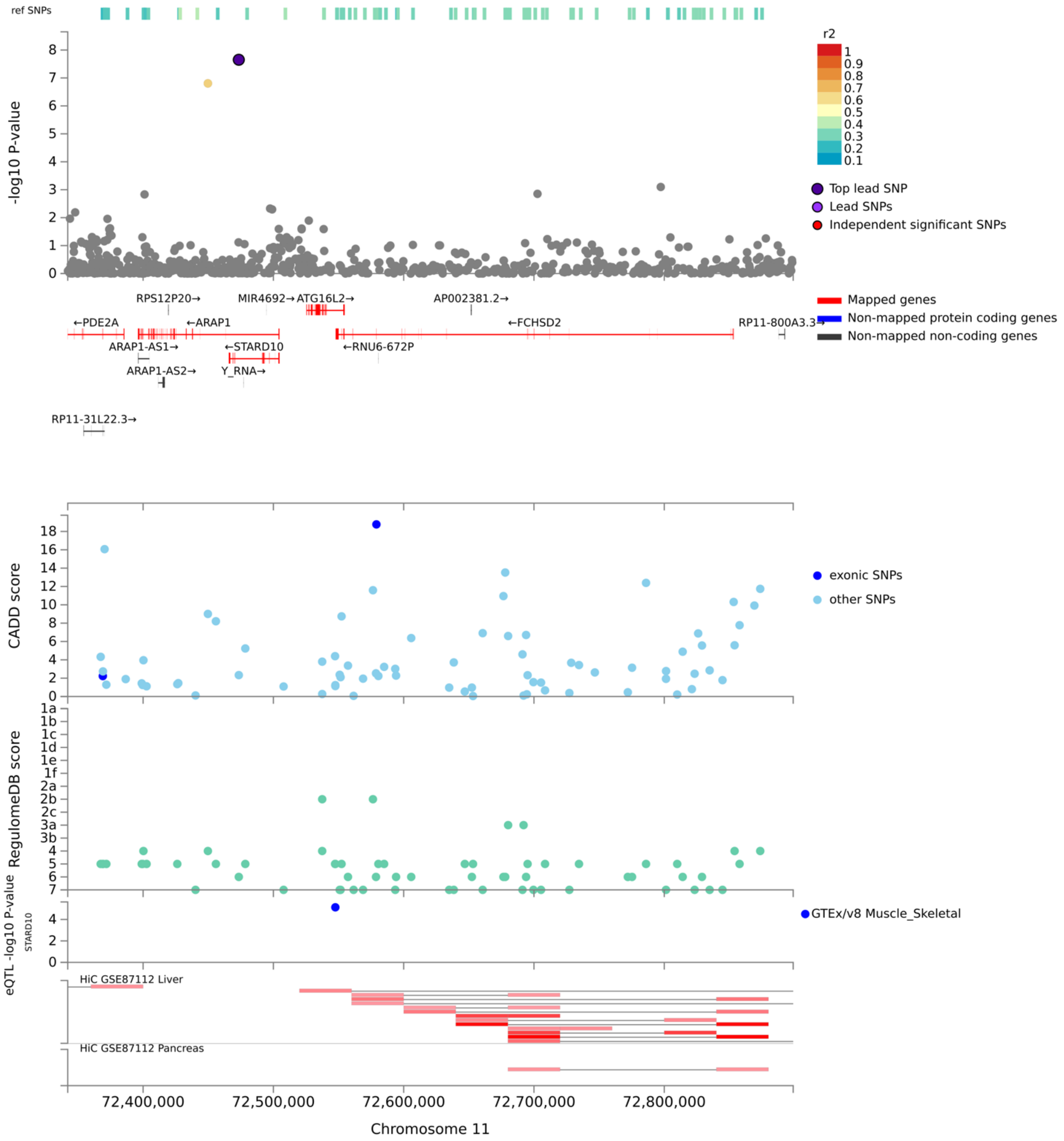

**Figure S14:** Regional plot of the **chr11q22.1** locus genome-wide significant in multivariate analysis of fructosamine and glycated albumin using BMI-unadjusted model on **sex-combined** genotyped/imputed data on **African ancestry** participants. The panels show GWAS association statistics, CADD score, Regulome DB score, eQTL and chromatin interaction information from FUMA. The *cis*-eQTL gene mapping used GTEx v8 data in T2D-relevant tissues and another data source on pancreatic islets. The 3D chromatin interaction gene mapping used Hi-C data in T2D-relevant tissues from GSE87112.

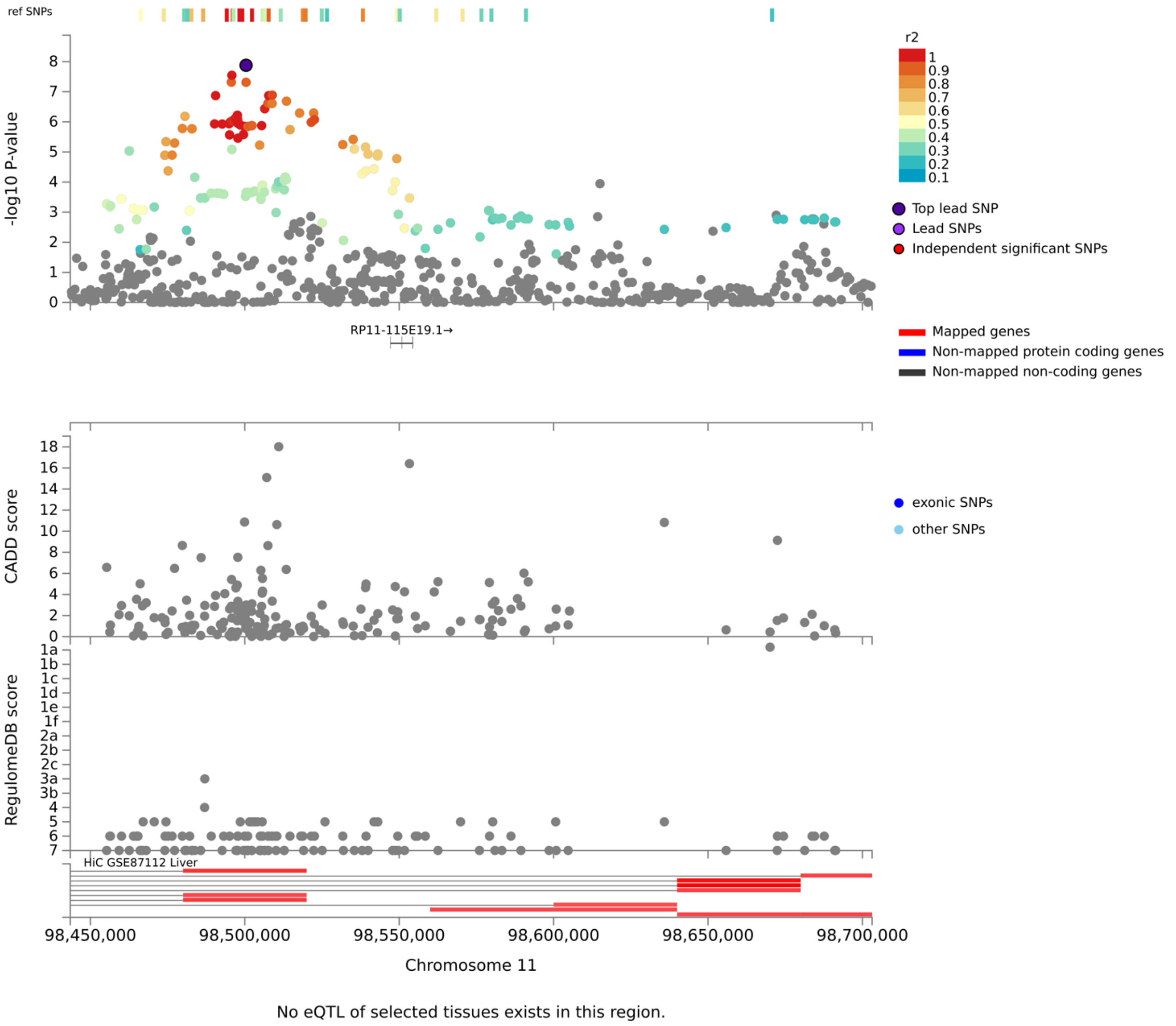

**Figure S15:** Regional plot of the **chr17q24.2** locus genome-wide significant in multivariate analysis of fructosamine and glycated albumin using BMI-unadjusted model on **sex-combined** genotyped/imputed data on **African ancestry** participants. The panels show GWAS association statistics, CADD score, Regulome DB score, eQTL and chromatin interaction information from FUMA. The *cis*-eQTL gene mapping used GTEx v8 data in T2D-relevant tissues and another data source on pancreatic islets. The 3D chromatin interaction gene mapping used Hi-C data in T2D-relevant tissues from GSE87112.

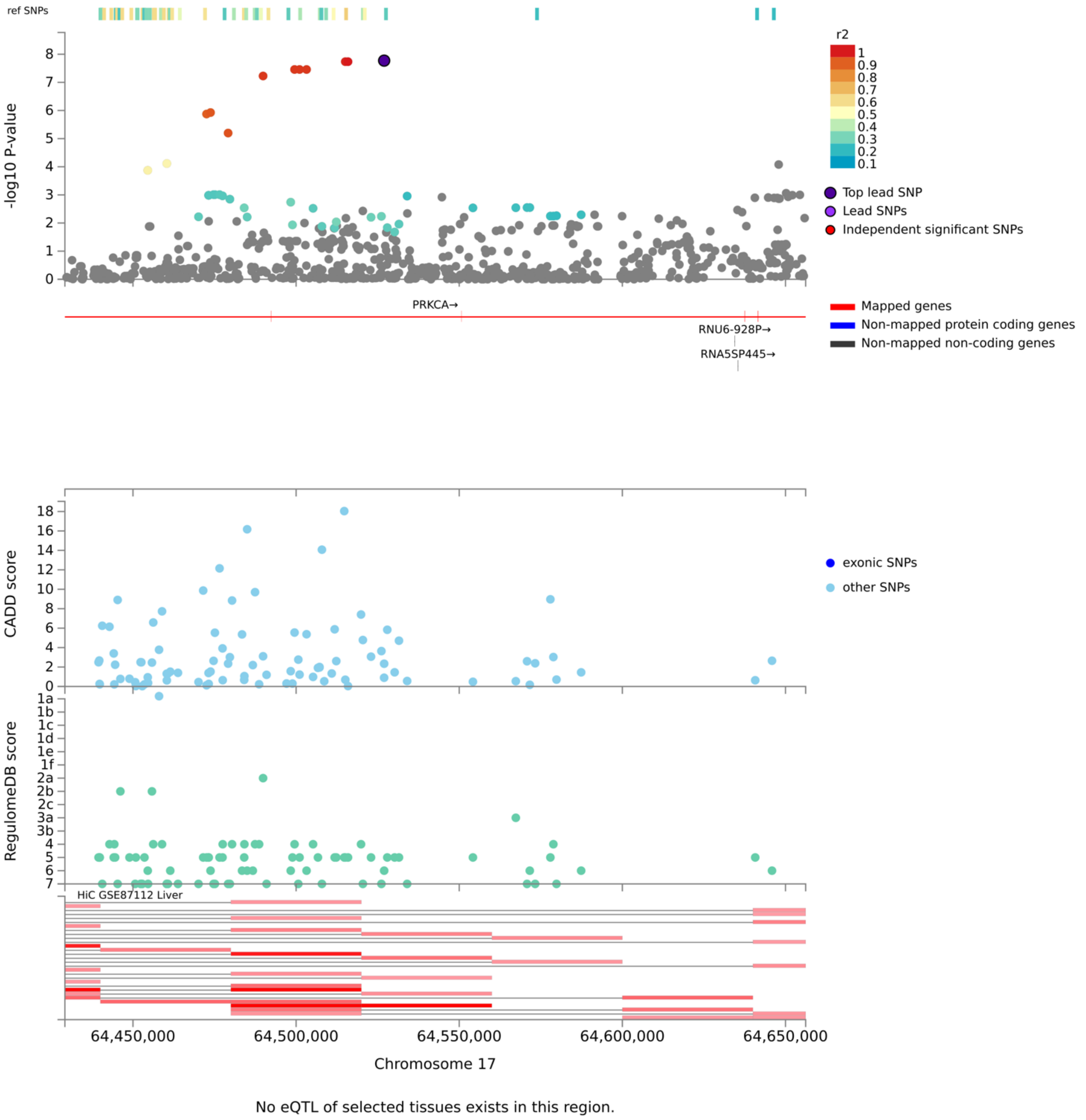

**Figure S16:** Manhattan plots of GAMuT association results from the gene burden multivariate analysis of fructosamine and glycated albumin using BMI-unadjusted model on sex-combined, multi-ancestry whole-exome sequence data.

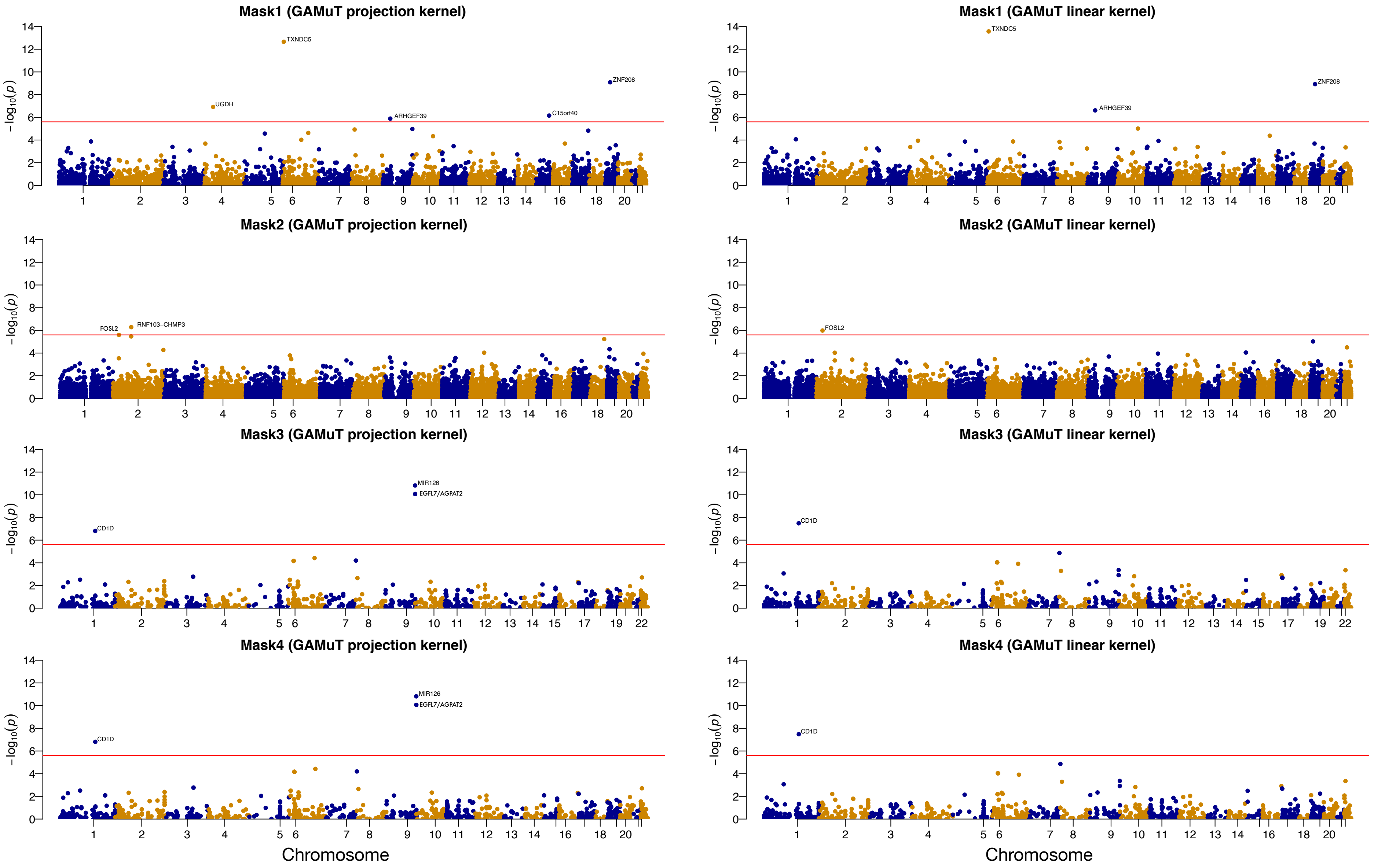

**Figure S17:** QQ plots of GAMuT association results from the gene burden multivariate analysis of fructosamine and glycated albumin using BMI-unadjusted model on sex-combined, multi-ancestry whole-exome sequence data.

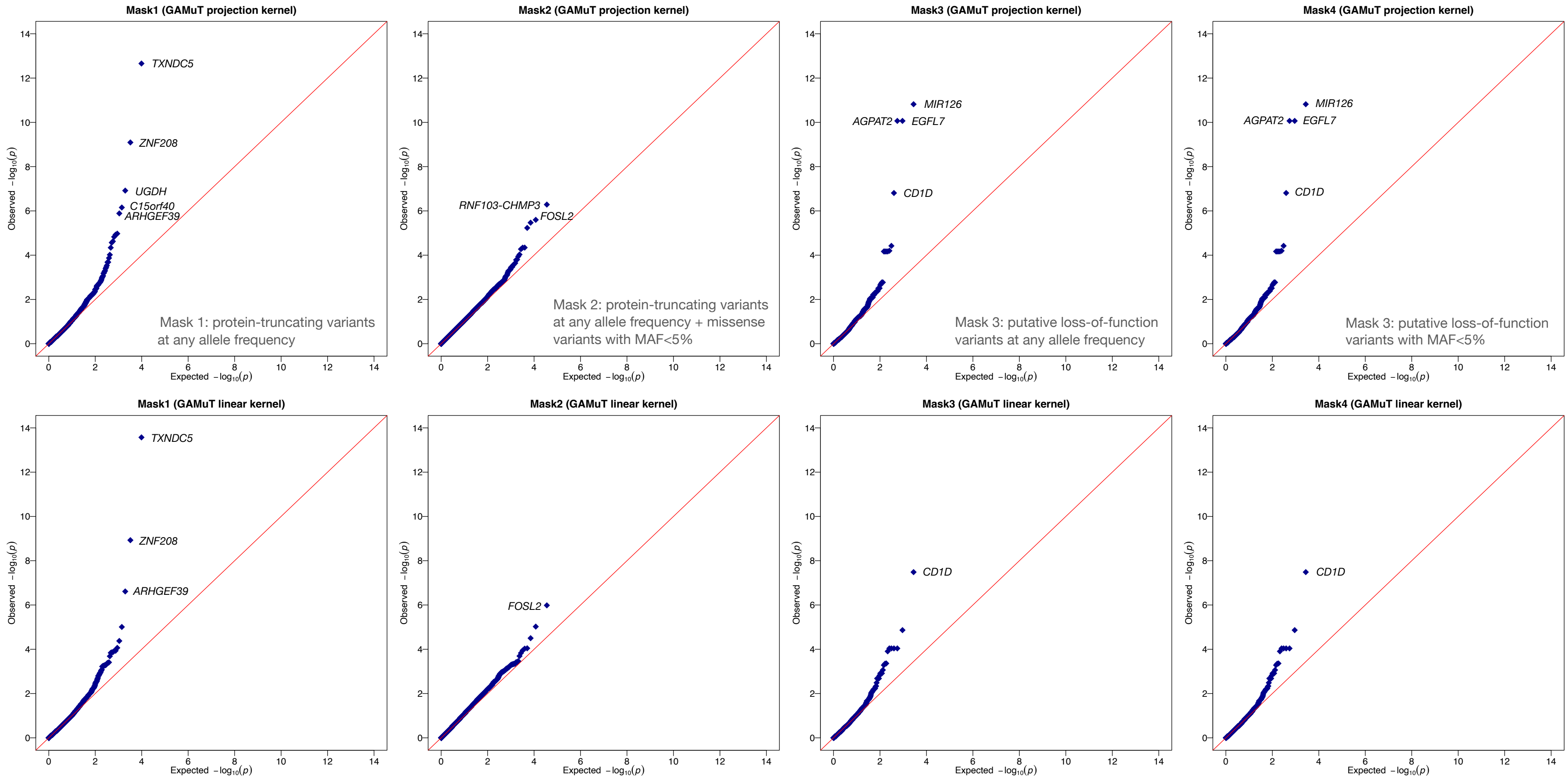

### SUPPLEMENTARY TABLES

**Table S1:** Demographic and clinical characteristics of study participants as used in multi-ancestry and ancestry-stratified analyses[1].

| | Genotyped sample | | | Exome sequenced sample | | |
|---|---|---|---|---|---|---|
| | European ancestry ($n = 7{,}359$) | African ancestry ($n = 2{,}004$) | Multi-ancestry ($n = 9{,}411$) | European ancestry ($n = 5{,}986$) | African ancestry ($n = 2{,}003$) | Multi-ancestry ($n = 8{,}898$) |
| Female | 3,992 (54%) | 1,258 (63%) | 5,275 (56%) | 3,294 (55%) | 1,258 (63%) | 5,099 (57%) |
| Age | 56.9 (5.7) | 55.9 (5.7) | 56.7 (5.7) | 57.0 (5.6) | 55.7 (5.7) | 56.7 (5.7) |
| Study center | | | | | | |
| Jackson, MS | 0 | 1,787 (89%) | 1,799 (19%) | 0 | 1,803 (90%) | 2,070 (23%) |
| Forsyth county, NC | 2,214 (30%) | 217 (11%) | 2,442 (26%) | 1,460 (24%) | 200 (10%) | 1,871 (21%) |
| Washington county, MD | 2,336 (32%) | 0 | 2,347 (25%) | 1,798 (30%) | 0 | 2,002 (22%) |
| Minneapolis, MN | 2,809 (38%) | 0 | 2,823 (30%) | 2,728 (46%) | 0 | 2,955 (33%) |
| Fructosamine (µmol/L) | 226.6 (22.9) | 238.5 (34.8) | 229.1 (26.3) | 226.7 (22.0) | 237.9 (30.3) | 229.8 (25.9) |
| Glycated albumin (%) | 12.6 (1.6) | 13.7 (2.6) | 12.8 (1.9) | 12.6 (1.5) | 13.6 (2.2) | 12.9 (1.9) |
| HbA1c (%)[2] | 5.4 (0.5) | 5.8 (0.8) | 5.5 (0.6) | 5.4 (0.5) | 5.8 (0.8) | 5.5 (0.6) |
| Fasting glucose (mg/dL)[3] | 103.9 (17.0) | 109.2 (24.8) | 105 (19.0) | 103.7 (16.7) | 109.0 (24.0) | 105.2 (20.0) |

[1]Continuous variables shown as mean (SD); categorical variables shown as counts (percentages)
[2]HbA1c *genotyped sample* European ancestry participants n=7,280, African ancestry participants n=1,965, Multi-ancestry participants n=9,291; *exome sequenced sample* European ancestry participants n=5,922, African ancestry participants n=1,967, Multi-ancestry participants n=8,790.
[3]Fasting glucose *genotyped sample* European ancestry participants n=7,263, African ancestry participants n=1,932, Multi-ancestry participants n=9,242; *exome sequenced sample* European ancestry participants n=5,913, African ancestry participants n=1,933, Multi-ancestry participants n=8,734.

**Table S2:** Pearson's correlations among biomarkers in multi-ancestry genotyped sample ($n = 9{,}411$) and in whole-exome sequenced sample ($n = 8{,}621$).

| | Fructosamine (µmol/L) | Glycated Albumin (%) | HbA1c (%) | Fasting Glucose (mg/dL) |
|---|---|---|---|---|
| Fructosamine (µmol/L) | 1 | | | |
| Glycated Albumin (%) | 0.79, 0.80 | 1 | | |
| HbA1c (%) | 0.52, 0.55 | 0.61, 0.64 | 1 | |
| Fasting Glucose (mg/dL) | 0.58, 0.60 | 0.61, 0.65 | 0.72, 0.74 | 1 |

**Table S3:** Replication of loci detected at suggestive threshold $10^{-6}$ from our common variant multi-phenotype analysis in one of the largest multi-ancestry GWAS of T2D from the Million Veteran Program (MVP) (Vojkovic et al, 2020[32]). Association p-values $< 10^{-3}$ in MVP are bold-faced.

| Locus | Nearest gene | rsID (lead SNP) | Position (hg19) | Effect allele | Fructosamine & Glycated albumin (ARIC) | T2D (European ancestry from MVP: 148,726 cases/965,732 controls) | | | T2D (African American from MVP: 24,646 cases/31,446 controls) | | | T2D (Multi-ancestry meta-analysis from MVP: 228,499 cases/1,178,783 controls) | | |
|---|---|---|---|---|---|---|---|---|---|---|---|---|---|---|
| | | | | | P-value* (metaUSAT) | Effect allele freq. | Effect size $\beta^{\S}$ | P-value* | Effect allele freq. | Effect size $\beta^{\S}$ | P-value* | Effect allele freq. | Effect size $\beta^{\S}$ | P-value* |
| **Multi-ancestry** | | | | | | | | | | | | | | |
| 1p32.2 | *RP1-158P9.1* | rs1331859 | 56594111 | A | $4.4 \times 10^{-7}$ | 0.349 | 0.010 | $4.1 \times 10^{-2}$ | 0.079 | 0.053 | $3.6 \times 10^{-2}$ | 0.317 | 0.011 | $7.3 \times 10^{-3}$ |
| 2q37.1 | *UGT1A1* | rs4148325 | 234673309 | T | $5.9 \times 10^{-7}$ | 0.322 | 0.026 | $\mathbf{8.9 \times 10^{-8}}$ | 0.446 | -0.002 | $9.0 \times 10^{-1}$ | 0.320 | 0.023 | $\mathbf{2.6 \times 10^{-8}}$ |
| 4q23 | *TSPAN5/ RP11-724M22.1* | rs75603568 | 99426458 | A | $9.3 \times 10^{-7}$ | 0.005 | 0.089 | $1.2 \times 10^{-1}$ | 0.122 | 0.028 | $2.1 \times 10^{-1}$ | 0.087 | 0.012 | $3.6 \times 10^{-1}$ |
| 7p21.2 | *ISPD* | rs1859423 | 16343897 | C | $7.0 \times 10^{-7}$ | 0.021 | 0.009 | $5.9 \times 10^{-1}$ | 0.081 | 0.026 | $3.0 \times 10^{-1}$ | 0.133 | 0.002 | $7.8 \times 10^{-1}$ |
| 11q13.4 | *ARAP1/ STARD10* | rs116714277 | 72473447 | T | $2.8 \times 10^{-8}$ | | | | 0.063 | 0.047 | $9.2 \times 10^{-2}$ | 0.060 | 0.042 | $1.3 \times 10^{-1}$ |
| 17q24.2 | *PRKCA* | rs59443763 | 64526988 | C | $1.4 \times 10^{-8}$ | | | | 0.067 | 0.008 | $7.5 \times 10^{-1}$ | 0.060 | 0.005 | $8.5 \times 10^{-1}$ |
| 19q13.33 | *FCGRT* | rs59774409 | 50016748 | T | $1.3 \times 10^{-7}$ | 0.084 | 0.020 | $1.9 \times 10^{-2}$ | 0.268 | 0.008 | $6.2 \times 10^{-1}$ | 0.115 | 0.025 | $\mathbf{3.7 \times 10^{-4}}$ |
| **European ancestry** | | | | | | | | | | | | | | |
| 1p31.3 | *ATG4C* | rs17316247 | 63258549 | C | $5.2 \times 10^{-7}$ | 0.280 | -0.010 | $1.5 \times 10^{-1}$ | | | | 0.260 | -0.012 | $6.5 \times 10^{-2}$ |
| 2p23.3 | *GCKR* | rs1260326 | 27730940 | T | $5.2 \times 10^{-8}$ | 0.403 | 0.064 | $\mathbf{2.6 \times 10^{-42}}$ | 0.141 | 0.034 | $6.3 \times 10^{-2}$ | 0.416 | 0.063 | $\mathbf{2.2 \times 10^{-57}}$ |
| 2q37.1 | *UGT1A1* | rs887829 | 234668570 | T | $2.2 \times 10^{-8}$ | 0328 | 0.027 | $\mathbf{3.4 \times 10^{-8}}$ | 0.446 | 0.001 | $9.4 \times 10^{-1}$ | 0.324 | 0.024 | $\mathbf{8.8 \times 10^{-9}}$ |
| 19q13.33 | *FCGRT* | rs59774409 | 50016748 | T | $2.4 \times 10^{-8}$ | 0.084 | 0.020 | $1.9 \times 10^{-2}$ | 0.268 | 0.008 | $6.2 \times 10^{-1}$ | 0.115 | 0.025 | $\mathbf{3.7 \times 10^{-4}}$ |
| **African ancestry** | | | | | | | | | | | | | | |
| 2p22.1 | *AC007317.1* | rs10490265 | 40979468 | T | $6.2 \times 10^{-8}$ | 0.362 | -0.001 | $7.7 \times 10^{-1}$ | 0.305 | 0.002 | $8.8 \times 10^{-1}$ | 0.328 | -0.005 | $2.5 \times 10^{-1}$ |
| 11q13.4 | *ARAP1/ STARD10* | rs116714277 | 72473447 | T | $2.2 \times 10^{-8}$ | | | | 0.063 | 0.047 | $9.2 \times 10^{-2}$ | 0.060 | 0.042 | $1.3 \times 10^{-1}$ |
| 11q22.1 | *RP11-115E19.1* | rs2438321 | 98500410 | G | $1.3 \times 10^{-8}$ | 0.268 | -0.004 | $4.0 \times 10^{-1}$ | 0.129 | -0.003 | $8.7 \times 10^{-1}$ | 0.275 | -0.003 | $4.6 \times 10^{-1}$ |
| 17q24.2 | *PRKCA* | rs59443763 | 64526988 | C | $1.7 \times 10^{-8}$ | | | | 0.067 | 0.008 | $7.5 \times 10^{-1}$ | 0.060 | 0.005 | $8.5 \times 10^{-1}$ |
| 21q22.3 | *PRDM15* | rs62214725 | 43298715 | T | $9.6 \times 10^{-7}$ | 0.291 | 0.001 | $9.2 \times 10^{-1}$ | | | | 0.292 | 0.001 | $8.2 \times 10^{-1}$ |

*BMI-unadjusted p-values are reported.

**Table S4:** Association results for the most significant SNPs of the loci identified from sex-stratified, multivariate analysis of fructosamine and glycated albumin using either BMI-unadjusted or adjusted model on multi-ancestry genotyped/imputed data. These loci were identified by metaUSAT at a suggestive threshold of $10^{-6}$. Bold-faced metaUSAT p-values indicate if a locus was identified from a BMI-unadjusted or a BMI-adjusted model. For a SNP, the metaUSAT p-value $< 5 \times 10^{-8}$ indicates its statistically significant association with at least one of fructosamine and glycated albumin at the genome-wide level, which may or may not be significant in single-phenotype analysis. The effect sizes and p-values for individual phenotypes are from the BMI-unadjusted sex-specific model only.

| Locus | Nearest gene | rsID (lead SNP) | Position (hg19) | Effect allele | Fructosamine | | Glycated Albumin | | Multivariate analysis (metaUSAT) | |
|---|---|---|---|---|---|---|---|---|---|---|
| | | | | | Effect size $\beta$§ | P-value | Effect size $\beta$§ | P-value | BMI-unadjusted p-value | BMI-adjusted p-value |
| **Multi-ancestry, female** | | | | | | | | | | |
| 4q25 | *DKK* | rs7680108 | 107933988 | C | 0.047 | $7.2 \times 10^{-6}$ | 0.065 | $1.0 \times 10^{-7}$ | $\mathbf{2.5 \times 10^{-7}}$ | $3.5 \times 10^{-6}$ |
| | *COL25A1* | rs115329747 | 109987836 | A | 0.059 | $3.6 \times 10^{-7}$ | 0.044 | $1.3 \times 10^{-3}$ | $2.1 \times 10^{-6}$ | $\mathbf{5.5 \times 10^{-7}}$ |
| 8p22 | *DLC1* | rs76178071 | 12930556 | T | 0.036 | $2.4 \times 10^{-6}$ | 0.045 | $6.5 \times 10^{-7}$ | $\mathbf{4.0 \times 10^{-7}}$ | $\mathbf{5.5 \times 10^{-8}}$ |
| 8q24.11 | *SLC30A8* | rs141866157 | 118206415 | T | 0.012 | $1.5 \times 10^{-5}$ | 0.017 | $1.0 \times 10^{-7}$ | $\mathbf{4.0 \times 10^{-7}}$ | $\mathbf{1.8 \times 10^{-7}}$ |
| 11p15.1 | *SOX6* | rs1401454 | 16250183 | T | -0.0087 | $8.6 \times 10^{-6}$ | -0.0026 | $2.6 \times 10^{-1}$ | $\mathbf{2.5 \times 10^{-7}}$ | $1.9 \times 10^{-6}$ |
| 11q13.4 | *ARAP1/ STARD10* | rs116714277 | 72473447 | T | 0.052 | $2.7 \times 10^{-7}$ | 0.061 | $3.3 \times 10^{-7}$ | $\mathbf{7.4 \times 10^{-8}}$ | $\mathbf{1.7 \times 10^{-7}}$ |
| 17q21.33 | *CA10* | rs73989066 | 50142984 | T | 0.016 | $1.0 \times 10^{-5}$ | 0.0056 | $2.0 \times 10^{-1}$ | $1.5 \times 10^{-6}$ | $\mathbf{7.0 \times 10^{-7}}$ |
| 19q13.33 | *RPS11* | rs2077300 | 50003253 | T | -0.013 | $2.5 \times 10^{-7}$ | -0.013 | $2.6 \times 10^{-5}$ | $\mathbf{9.5 \times 10^{-7}}$ | $\mathbf{8.7 \times 10^{-7}}$ |
| 19q13.41 | *SIGLEC14* | rs116588425 | 52165773 | T | 0.035 | $2.0 \times 10^{-4}$ | 0.056 | $4.5 \times 10^{-7}$ | $2.7 \times 10^{-6}$ | $\mathbf{6.6 \times 10^{-7}}$ |
| **Multi-ancestry, male** | | | | | | | | | | |
| 3p14.1 | *PSMD6-AS2* | rs73834175 | 63995489 | G | 0.054 | $5.2 \times 10^{-7}$ | 0.056 | $1.1 \times 10^{-5}$ | $\mathbf{8.7 \times 10^{-7}}$ | $5.6 \times 10^{-6}$ |
| 3p13 | *SHQ1* | rs61287719 | 72721724 | T | 0.057 | $3.5 \times 10^{-7}$ | 0.060 | $7.1 \times 10^{-6}$ | $\mathbf{5.5 \times 10^{-7}}$ | $3.9 \times 10^{-6}$ |
| 4q22.3 | *UNC5C* | rs78307336 | 96261643 | T | 0.048 | $1.5 \times 10^{-5}$ | 0.065 | $5.9 \times 10^{-7}$ | $1.2 \times 10^{-6}$ | $\mathbf{3.5 \times 10^{-7}}$ |
| 5p15.1 | *FTH1P10* | rs2650945 | 17351097 | T | 0.019 | $3.9 \times 10^{-7}$ | 0.018 | $3.5 \times 10^{-5}$ | $1.7 \times 10^{-6}$ | $\mathbf{9.5 \times 10^{-7}}$ |
| 5q32 | *GRXCR2* | rs73308169 | 145267978 | A | 0.067 | $1.9 \times 10^{-6}$ | 0.077 | $3.4 \times 10^{-6}$ | $\mathbf{9.9 \times 10^{-7}}$ | $\mathbf{2.9 \times 10^{-7}}$ |
| 8q21.13 | *STMN2* | rs113747263 | 80573416 | A | 0.060 | $4.1 \times 10^{-6}$ | 0.074 | $1.8 \times 10^{-6}$ | $1.1 \times 10^{-6}$ | $\mathbf{2.5 \times 10^{-7}}$ |
| 10p11.23 | *KIAA1462* | rs16930899 | 30261818 | T | 0.061 | $2.4 \times 10^{-6}$ | 0.067 | $1.1 \times 10^{-5}$ | $2.2 \times 10^{-6}$ | $\mathbf{8.5 \times 10^{-7}}$ |
| 11p14.3 | *RP11-266A24.1* | rs181527286 | 23215185 | T | 0.079 | $2.9 \times 10^{-7}$ | 0.089 | $8.7 \times 10^{-7}$ | $\mathbf{1.5 \times 10^{-7}}$ | $\mathbf{4.8 \times 10^{-8}}$ |
| 12q24.32 | *RP11-575F12.1* | rs1732470 | 127446253 | T | 0.045 | $4.2 \times 10^{-7}$ | 0.032 | $2.2 \times 10^{-3}$ | $1.7 \times 10^{-6}$ | $\mathbf{7.9 \times 10^{-7}}$ |
| 13q34 | *TEX29* | rs79276590 | 112106288 | C | 0.054 | $1.4 \times 10^{-4}$ | 0.097 | $6.7 \times 10^{-9}$ | $\mathbf{3.0 \times 10^{-8}}$ | $\mathbf{9.0 \times 10^{-7}}$ |
| 14q24.1 | *ACTN1* | rs2268973 | 69424022 | T | 0.0048 | $6.0 \times 10^{-2}$ | 0.014 | $2.2 \times 10^{-6}$ | $\mathbf{7.9 \times 10^{-7}}$ | $1.1 \times 10^{-6}$ |
| 17q25.1 | *POLR3KP2* | rs12939639 | 71138612 | T | 0.059 | $4.0 \times 10^{-6}$ | 0.068 | $7.4 \times 10^{-6}$ | $2.3 \times 10^{-6}$ | $\mathbf{4.6 \times 10^{-7}}$ |

§$\beta$ denotes mean change in log(outcome) for every additional copy of the effect allele.

**Table S5:** Characterization of detected loci from common variant multi-phenotype analysis as "glycemic", "maybe glycemic" vs "non-glycemic". The percentage change in effect size column indicates $100 \times (\beta_{\text{no-FG-adj}} - \beta_{\text{FG-adj}})/\beta_{\text{no-FG-adj}}$, where a positive value of 25% or more (bold-faced) is indicative of a potential glycemic pathway mediated by fasting glucose (FG). While FG-adjusted effect size estimates ($\beta_{\text{FG-adj}}$) for fructosamine and glycated albumin are provided in this table, the estimates $\beta_{\text{no-FG-adj}}$ for both traits are provided in Table 1. Fasting glucose and HbA1c p-values $<$ 0.005 are also bold-faced. The final column indicates classification based on details in Table S7.

| Locus | Nearest gene | rsID (lead SNP) | Position (hg19) | Effect allele | Fructosamine (FG-adjusted) | | | Glycated Albumin (FG-adjusted) | | | Fasting glucose (FG) | | HbA1c | | Classified as glycemic? |
|---|---|---|---|---|---|---|---|---|---|---|---|---|---|---|---|
| | | | | | Effect size $\beta^{\S}$ | % change in effect size | P-value | Effect size $\beta^{\S}$ | % change in effect size | P-value | Effect size $\beta^{\S}$ | P-value | Effect size $\beta^{\S}$ | P-value | |
| **Multi-ancestry** | | | | | | | | | | | | | | | |
| 1p32.2 | *RP1-158P9.1* | rs1331859 | 56594111 | A | 0.00073 | -318.7% | $6.1 \times 10^{-1}$ | -0.0058 | 13.0% | $5.6 \times 10^{-4}$ | -0.0019 | $3.9 \times 10^{-1}$ | -0.0017 | $2.5 \times 10^{-1}$ | No |
| 2q37.1 | *UGT1A1* | rs4148325 | 234673309 | T | 0.0054 | -13.8% | $3.3 \times 10^{-5}$ | -0.00049 | **49.7%** | $7.5 \times 10^{-1}$ | 0.00019 | $9.3 \times 10^{-1}$ | 0.00020 | $8.9 \times 10^{-1}$ | No |
| 4q23 | *TSPAN5/ RP11-724M22.1* | rs75603568 | 99426458 | A | 0.013 | **55.0%** | $7.9 \times 10^{-5}$ | 0.016 | **54.3%** | $4.3 \times 10^{-3}$ | 0.011 | $2.5 \times 10^{-1}$ | 0.014 | $3.0 \times 10^{-2}$ | Glycemic |
| 7p21.2 | *ISPD* | rs1859423 | 16343897 | C | 0.021 | **37.3%** | $1.9 \times 10^{-4}$ | 0.027 | **34.2%** | $2.4 \times 10^{-5}$ | 0.024 | $1.9 \times 10^{-2}$ | 0.019 | $9.7 \times 10^{-3}$ | Glycemic |
| 11q13.4 | *ARAP1/ STARD10* | rs116714277 | 72473447 | T | 0.024 | **45.0%** | $4.0 \times 10^{-4}$ | 0.030 | **45.7%** | $1.5 \times 10^{-4}$ | 0.050 | $\mathbf{8.3 \times 10^{-5}}$ | 0.038 | $\mathbf{1.6 \times 10^{-5}}$ | Glycemic |
| 17q24.2 | *PRKCA* | rs59443763 | 64526988 | C | 0.017 | **54.4%** | $5.5 \times 10^{-3}$ | 0.029 | **46.1%** | $3.1 \times 10^{-5}$ | 0.040 | $\mathbf{4.0 \times 10^{-4}}$ | 0.024 | $\mathbf{2.6 \times 10^{-3}}$ | Glycemic |
| 19q13.33 | *FCGRT* | rs59774409 | 50016748 | T | -0.018 | -3.0% | $1.0 \times 10^{-8}$ | -0.021 | 0.3% | $1.3 \times 10^{-8}$ | 0.0052 | $2.9 \times 10^{-1}$ | 0.00039 | $9.0 \times 10^{-1}$ | Glycemic |
| **European ancestry** | | | | | | | | | | | | | | | |
| 1p31.3 | *ATG4C* | rs17316247 | 63258549 | C | 0.0063 | 23.8% | $4.9 \times 10^{-5}$ | 0.0071 | **27.9%** | $1.3 \times 10^{-4}$ | 0.0049 | $4.5 \times 10^{-2}$ | 0.0047 | $\mathbf{2.9 \times 10^{-3}}$ | Maybe |
| 2p23.3 | *GCKR* | rs1260326 | 27730940 | T | -0.00093 | **74.9%** | $5.1 \times 10^{-1}$ | -0.0067 | **34.3%** | $5.2 \times 10^{-5}$ | -0.0097 | $\mathbf{7.8 \times 10^{-6}}$ | -0.0027 | $6.4 \times 10^{-2}$ | Glycemic |
| 2q37.1 | *UGT1A1* | rs887829 | 234668570 | T | 0.0071 | 4.9% | $1.4 \times 10^{-6}$ | 0.00005 | **93.2%** | $9.8 \times 10^{-1}$ | 0.0016 | $4.9 \times 10^{-1}$ | 0.0011 | $4.5 \times 10^{-1}$ | No |
| 19q13.33 | *FCGRT* | rs59774409 | 50016748 | T | -0.019 | -3.4% | $2.0 \times 10^{-9}$ | -0.022 | 0.1% | $3.9 \times 10^{-9}$ | 0.0055 | $2.6 \times 10^{-1}$ | 0.00007 | $9.8 \times 10^{-1}$ | Glycemic |
| **African ancestry** | | | | | | | | | | | | | | | |
| 2p22.1 | *AC007317.1* | rs10490265 | 40979468 | T | 0.020 | **36.6%** | $3.5 \times 10^{-6}$ | 0.013 | **52.0%** | $8.6 \times 10^{-3}$ | 0.0089 | $2.4 \times 10^{-1}$ | 0.0038 | $4.8 \times 10^{-1}$ | Glycemic |
| 11q13.4 | *ARAP1/ STARD10* | rs116714277 | 72473447 | T | 0.024 | **46.3%** | $3.9 \times 10^{-4}$ | 0.029 | **47.9%** | $1.5 \times 10^{-4}$ | 0.048 | $\mathbf{1.6 \times 10^{-4}}$ | 0.037 | $\mathbf{2.3 \times 10^{-5}}$ | Glycemic |
| 11q22.1 | *RP11-115E19.1* | rs2438321 | 98500410 | G | 0.024 | **37.0%** | $9.7 \times 10^{-7}$ | 0.016 | **52.7%** | $5.4 \times 10^{-3}$ | 0.029 | $\mathbf{2.5 \times 10^{-3}}$ | 0.020 | $\mathbf{1.8 \times 10^{-3}}$ | Glycemic |
| 17q24.2 | *PRKCA* | rs59443763 | 64526988 | C | 0.015 | **60.4%** | $1.3 \times 10^{-2}$ | 0.025 | **53.2%** | $2.4 \times 10^{-4}$ | 0.042 | $\mathbf{1.6 \times 10^{-4}}$ | 0.024 | $\mathbf{1.8 \times 10^{-3}}$ | Glycemic |
| 21q22.3 | *PRDM15* | rs62214725 | 43298715 | T | 0.0088 | **69.1%** | $8.8 \times 10^{-2}$ | 0.017 | **56.7%** | $2.7 \times 10^{-3}$ | 0.042 | $\mathbf{2.4 \times 10^{-5}}$ | 0.013 | $5.9 \times 10^{-2}$ | Glycemic |

**Table S6:** Characterization of detected genes from rare variant set-based multi-phenotype analysis of sex-combined multi-ancestry data as "glycemic", "maybe glycemic" vs "non-glycemic". Fasting glucose and HbA1c p-values $< 0.005$ are bold-faced. Note, projection and linear kernels were used to summarize multi-phenotype information and thus multi-phenotype analyses have different GAMuT p-values depending on kernel choice (unlike single-phenotype analyses). Individuals contributing to MAC for each trait can be slightly different depending on which individuals have measurements on some or all these traits. The final column indicates classification based on details in Table S9.

| Chr | Gene | No. of variants | Fructosamine & Glycated albumin | | | Fructosamine | | Glycated albumin | | Fasting glucose | | HbA1c | | Classified as glycemic? |
|---|---|---|---|---|---|---|---|---|---|---|---|---|---|---|
| | | | Total MAC | GAMuT p-value | | Total MAC | GAMuT p-value | Total MAC | GAMuT p-value | Total MAC | GAMuT p-value | Total MAC | GAMuT p-value | |
| | | | | Projection kernel | Linear kernel | | | | | | | | | |
| **Mask 1:** protein-truncating variants at any allele frequency | | | | | | | | | | | | | | |
| 4 | *UGDH* | 3 | 5 | $1.2 \times 10^{-7}$ | $1.2 \times 10^{-4}$ | 5 | $1.2 \times 10^{-6}$ | 5 | $1.3 \times 10^{-1}$ | 5 | $7.0 \times 10^{-1}$ | 5 | $9.9 \times 10^{-1}$ | No |
| 6 | *TXNDC5* | 8 | 5 | $2.2 \times 10^{-13}$ | $2.7 \times 10^{-14}$ | 5 | $7.4 \times 10^{-13}$ | 5 | $2.6 \times 10^{-12}$ | 4 | $\mathbf{7.8 \times 10^{-11}}$ | 4 | $\mathbf{2.7 \times 10^{-11}}$ | Glycemic |
| 9 | *ARHGEF39* | 9 | 6 | $1.3 \times 10^{-6}$ | $2.4 \times 10^{-7}$ | 6 | $4.4 \times 10^{-6}$ | 6 | $5.1 \times 10^{-7}$ | 6 | $\mathbf{2.9 \times 10^{-6}}$ | 6 | $\mathbf{8.6 \times 10^{-5}}$ | Glycemic |
| 15 | *C15orf40* | 3 | 5 | $7.0 \times 10^{-7}$ | $5.3 \times 10^{-3}$ | 5 | $2.2 \times 10^{-4}$ | 5 | $9.8 \times 10^{-1}$ | 4 | $3.3 \times 10^{-1}$ | 4 | $8.5 \times 10^{-1}$ | No |
| 19 | *ZNF208* | 6 | 7 | $8.1 \times 10^{-10}$ | $1.2 \times 10^{-9}$ | 7 | $2.2 \times 10^{-6}$ | 7 | $8.1 \times 10^{-11}$ | 7 | $\mathbf{1.3 \times 10^{-10}}$ | 7 | $\mathbf{5.1 \times 10^{-8}}$ | Glycemic |
| **Mask 2:** protein-truncating variants at any allele frequency + missense variants with MAF <5% | | | | | | | | | | | | | | |
| 2 | *FOSL2* | 34 | 60 | $2.5 \times 10^{-6}$ | $1.0 \times 10^{-6}$ | 60 | $4.3 \times 10^{-7}$ | 60 | $6.9 \times 10^{-5}$ | 58 | $\mathbf{6.8 \times 10^{-4}}$ | 58 | $1.3 \times 10^{-2}$ | Glycemic |
| 2 | *RNF103-CHMP3* | 23 | 36 | $5.2 \times 10^{-7}$ | $9.3 \times 10^{-5}$ | 36 | $5.5 \times 10^{-2}$ | 36 | $1.5 \times 10^{-6}$ | 36 | $1.4 \times 10^{-2}$ | 36 | $\mathbf{7.9 \times 10^{-6}}$ | Maybe |
| **Mask 3:** putative loss-of-function variants at any allele frequency | | | | | | | | | | | | | | |
| 1 | *CD1D* | 5 | 9 | $1.6 \times 10^{-7}$ | $3.3 \times 10^{-8}$ | 9 | $4.8 \times 10^{-8}$ | 9 | $1.5 \times 10^{-6}$ | 9 | $\mathbf{2.2 \times 10^{-6}}$ | 9 | $\mathbf{1.8 \times 10^{-5}}$ | Glycemic |
| 9 | *EGFL7* | 8 | 7 | $8.5 \times 10^{-11}$ | $1.2 \times 10^{-3}$ | 7 | $2.4 \times 10^{-5}$ | 7 | $4.8 \times 10^{-1}$ | 5 | $1.2 \times 10^{-1}$ | 5 | $4.9 \times 10^{-1}$ | No |
| 9 | *MIR126* | 7 | 6 | $1.5 \times 10^{-11}$ | $4.3 \times 10^{-4}$ | 6 | $3.6 \times 10^{-6}$ | 6 | $6.7 \times 10^{-1}$ | 4 | $3.1 \times 10^{-1}$ | 4 | $8.5 \times 10^{-1}$ | No |
| 9 | *AGPAT2* | 8 | 7 | $8.5 \times 10^{-11}$ | $1.2 \times 10^{-3}$ | 7 | $2.4 \times 10^{-5}$ | 7 | $4.8 \times 10^{-1}$ | 5 | $1.2 \times 10^{-1}$ | 5 | $4.9 \times 10^{-1}$ | Maybe |
| **Mask 4:** putative loss-of-function variants with MAF <5% | | | | | | | | | | | | | | |
| 1 | *CD1D* | 5 | 9 | $1.6 \times 10^{-7}$ | $3.3 \times 10^{-8}$ | 9 | $4.8 \times 10^{-8}$ | 9 | $1.5 \times 10^{-6}$ | 9 | $\mathbf{2.2 \times 10^{-6}}$ | 9 | $\mathbf{1.8 \times 10^{-5}}$ | Glycemic |
| 9 | *EGFL7* | 8 | 7 | $8.5 \times 10^{-11}$ | $1.2 \times 10^{-3}$ | 7 | $2.4 \times 10^{-5}$ | 7 | $4.8 \times 10^{-1}$ | 5 | $1.2 \times 10^{-1}$ | 5 | $4.9 \times 10^{-1}$ | No |
| 9 | *MIR126* | 7 | 6 | $1.5 \times 10^{-11}$ | $4.3 \times 10^{-4}$ | 6 | $3.6 \times 10^{-6}$ | 6 | $6.7 \times 10^{-1}$ | 4 | $3.1 \times 10^{-1}$ | 4 | $8.5 \times 10^{-1}$ | No |
| 9 | *AGPAT2* | 8 | 7 | $8.5 \times 10^{-11}$ | $1.2 \times 10^{-3}$ | 7 | $2.4 \times 10^{-5}$ | 7 | $4.8 \times 10^{-1}$ | 5 | $1.2 \times 10^{-1}$ | 5 | $4.9 \times 10^{-1}$ | Maybe |

**Table S7:** Characterizing lead variants of significant loci from common variant analysis as "glycemic", "maybe glycemic" or "non-glycemic".

| Locus | rsID (lead SNP) | Condition 1 satisfied? | Condition 2 satisfied? | Condition 3 satisfied? | Condition 4 satisfied? | Condition 5 satisfied? | Classification |
|---|---|---|---|---|---|---|---|
| **Multi-ancestry** | | | | | | | |
| 1p32.2 | rs1331859 | No | No | No | No | No | Non-glycemic |
| 2q37.1 | rs4148325 | No | No | No | No | No | Non-glycemic |
| 4q23 | rs75603568 | Yes | No | No | No | No | Glycemic |
| 7p21.2 | rs1859423 | Yes | No | No | No | No | Glycemic |
| 11q13.4 | rs116714277 | Yes | No | Yes | No | Yes | Glycemic |
| 17q24.2 | rs59443763 | Yes | No | Yes | No | Yes | Glycemic |
| 19q13.33 | rs59774409 | No | Yes | No | No | No | Glycemic |
| **European ancestry** | | | | | | | |
| 1p31.3 | rs17316247 | No | No | No | No | Yes | Maybe glycemic |
| 2p23.3 | rs1260326 | Yes | Yes | Yes | Yes | No | Glycemic |
| 2q37.1 | rs887829 | No | No | No | No | No | Non-glycemic |
| 19q13.33 | rs59774409 | No | Yes | No | No | No | Glycemic |
| **African ancestry** | | | | | | | |
| 2p22.1 | rs10490265 | Yes | No | No | No | No | Glycemic |
| 11q13.4 | rs116714277 | Yes | No | Yes | No | Yes | Glycemic |
| 11q22.1 | rs2438321 | Yes | No | Yes | No | Yes | Glycemic |
| 17q24.2 | rs59443763 | Yes | No | Yes | No | Yes | Glycemic |
| 21q22.3 | rs62214725 | Yes | No | Yes | No | No | Glycemic |

**Condition 1:** lead variant effect size estimates for both fructosamine and glycated albumin are reduced by at least 25% when fasting glucose is adjusted in the model for ARIC
**Condition 2:** lead variant is associated ($p < 10^{-4}$) with any glycemic trait (except HbA1c) in a phenome-wide association (PheWAS) analysis of all traits available in Common Metabolic Diseases Knowledge Portal (CMD-KP) as of July 22, 2022 (updated information may be obtained from CMD-KP links provided in Table S8)
**Condition 3:** lead variant is associated with fasting glucose ($p < 0.005$) in ARIC
**Condition 4:** lead variant is associated ($p < 10^{-4}$) with HbA1c in a PheWAS of all traits available in CMD-KP as of July 22, 2022
**Condition 5:** lead variant is associated with HbA1c ($p < 0.005$) in ARIC

**Table S8:** Summary of information on significant loci (common variants) from Common Metabolic Diseases Knowledge Portal (as of July 22, 2022).

| Locus | rsID (lead SNP) | Information on lead SNP | Information on ±50 Kb region around lead SNP |
|---|---|---|---|
| **Multi-ancestry** | | | |
| 1p32.2 | rs1331859 | Implicated in genetics of cardiovascular, hematological and renal traits; Predicted regulatory regions that overlap the position of this variant are binding sites in tissues from skin of body, prostate gland, mammary gland https://md.hugeamp.org/variant.html?variant=rs1331859 | Implicated in genetics of lipids, cardiovascular, and hematological traits https://md.hugeamp.org/region.html?chr=1&end=56669112&phenotype=LDL&start=56519112 |
| 2q37.1 | rs4148325 | Implicated in genetics of bilirubin and lipids; Predicted regulatory regions that overlap the position of this variant include accessible chromatins (liver, adipose), enhancers (adipose), promoters (liver, muscle structure), binding sites etc from multiple tissues https://md.hugeamp.org/variant.html?variant=rs4148325 | Implicated in genetics of bilirubin, lipids and platelet count https://md.hugeamp.org/region.html?chr=2&end=234723309&phenotype=BILIRUBIN&start=234623309 |
| 4q23 | rs75603568 | – Predicted regulatory region that overlaps the position of this variant is accessible chromatin in muscle structure https://md.hugeamp.org/variant.html?variant=rs75603568 | Implicated in genetics of 2-hour glucose (BMI-adj), lipids, and alkaline phosphatase https://md.hugeamp.org/region.html?chr=4&end=99476458&phenotype=ALP&start=99376458 |
| 7p21.2 | rs1859423 | – Predicted regulatory region that overlaps the position of this variant is accessible chromatin in muscle structure https://md.hugeamp.org/variant.html?variant=rs1859423 | Implicated in genetics of calcium and HDL https://md.hugeamp.org/region.html?chr=7&end=16393897&phenotype=Ca&start=16293897 |
| 11q13.4 | rs116714277 | – Predicted regulatory regions that overlap the position of this variant are enhancers (pancreas, liver & blood), and binding sites in other tissues https://md.hugeamp.org/variant.html?variant=rs116714277 | Implicated in genetics of several glycemic traits (including T2D), hematological traits, and hepatic traits https://hugeamp.org/region.html?chr=11&end=72523447&phenotype=BasoCount&start=72423447 |
| 17q24.2 | rs59443763 | – Predicted regulatory regions that overlap the position of this variant are enhancers (CNS, stem cell), accessible chromatins (muscle structure, Cardiovascular system) and binding sites (stem cell) https://md.hugeamp.org/variant.html?variant=rs59443763 | Implicated in genetics of lipids, hepatic and cardiovascular traits, and platelet count https://hugeamp.org/region.html?chr=17&end=64576988&phenotype=LDL&start=64476988 |
| 19q13.33 | rs59774409 | Implicated in genetics of lipids, hepatic, and hematological traits; Predicted regulatory regions that overlap the position of this variant are promoters (adipose, liver, muscle structure, pancreas), accessible chromatins (adipose, liver, muscle structure, pancreas), binding sites (adipose, liver, muscle structure, pancreas) https://md.hugeamp.org/variant.html?variant=rs59774409 | Implicated in genetics of lipids, hepatic, and hematological traits https://hugeamp.org/region.html?chr=19&end=50066748&phenotype=CHOL&start=49966748 |
| **European ancestry** | | | |
| 1p31.3 | rs17316247 | – Predicted regulatory regions that overlap the position of this variant are promoters, accessible chromatins, binding sites in several tissues https://md.hugeamp.org/variant.html?variant=rs17316247 | Implicated in genetics of lipids, and hematological traits https://hugeamp.org/region.html?chr=1&end=63308549&phenotype=TG&start=63208549 |
| 2p23.3 | rs1260326 | Implicated in genetics of various lipids, glycemic traits (including T2D) as well as hematological traits; | Implicated in genetics of several glycemic traits (including T2D), lipids and hematological traits https://hugeamp.org/region.html?chr=2&end=27780940&phenotype=TG&start=27680940 |

| | | | |
|---|---|---|---|
| | | Predicted regulatory regions that overlap the position of this variant are promoter (pancreas), accessible chromatins (muscle structure, liver), enhancer (liver), binding sites etc. https://md.hugeamp.org/variant.html?variant=rs1260326 | |
| 2q37.1 | rs887829 | Implicated in genetics of bilirubin and lipids; Predicted regulatory regions that overlap the position of this variant are enhancers (liver), binding sites, accessible chromatins https://md.hugeamp.org/variant.html?variant=rs887829 | See above. |
| 19q13.3 3 | rs59774409 | See above. | See above. |
| **African ancestry** | | | |
| 2p22.1 | rs10490265 | – Predicted regulatory regions that overlap the position of this variant are enhancers (muscle structure, adipose), accessible chromatin https://md.hugeamp.org/variant.html?variant=rs10490265 | Implicated in genetics of blood pressure traits; suggestively significant for diabetic retinopathy https://hugeamp.org/region.html?chr=2&end=41029468&phenotype=PRI&start=40929468 |
| 11q13.4 | rs11671427 7 | See above. | See above. |
| 11q22.1 | rs2438321 | – https://md.hugeamp.org/variant.html?variant=rs2438321 | Suggestively implicated in genetics of hemorrhoids https://hugeamp.org/region.html?chr=11&end=98550410&phenotype=Hemorrhoids&start=98450410 |
| 17q24.2 | rs59443763 | See above. | See above. |
| 21q22.3 | rs62214725 | – Predicted regulatory regions that overlap the position of this variant are promoters (pancreas, adipose), binding sites in several tissues https://md.hugeamp.org/variant.html?variant=rs62214725 | Implicated in genetics of hematological traits https://hugeamp.org/region.html?chr=21&end=43348715&phenotype=PlatCount&start=43248715 |
| **Multi-ancestry, Males only** | | | |
| 13q34 | rs79276590 | – Predicted regulatory region that overlaps the position of this variant is accessible chromatin in muscle structure https://md.hugeamp.org/variant.html?variant=rs75603568 | Suggestively implicated in genetics of end-stage renal disease (ESRD) vs no ESRD adjusting for HbA1c-BMI https://md.hugeamp.org/region.html?chr=13&end=112156288&phenotype=ESRDvNonESRDadjHbA1cBMI&start=112056288 |

**Table S9:** Characterizing significant genes from set-based analyses of rare variants as “glycemic”, “maybe glycemic” or “non-glycemic”.

| **Gene** | **Condition 1 satisfied?** | **Condition 2 satisfied?** | **Condition 3 satisfied?** | **Condition 4 satisfied?** | **Classification** |
|---|---|---|---|---|---|
| **Mask 1** | | | | | |
| *UGDH* | No | No | No | No | Non-glycemic |
| *TXNDC5* | No | Yes | No | Yes | Glycemic |
| *ARHGEF39* | No | Yes | No | Yes | Glycemic |
| *C15orf40* | No | No | No | No | Non-glycemic |
| *ZNF208* | Yes | Yes | No | Yes | Glycemic |
| **Mask 2** | | | | | |
| *FOSL2* | No | Yes | No | No | Glycemic |
| *RNF103-CHMP3* | No | No | No | Yes | Maybe glycemic |
| **Mask 3 (also Mask 4)** | | | | | |
| *CD1D* | No | Yes | No | Yes | Glycemic |
| *EGFL7* | No | No | No | No | Non-glycemic |
| *MIR126* | No | No | No | No | Non-glycemic |
| *AGPAT2* | No | No | Yes | No | Maybe glycemic |

**Condition 1:** gene is associated ($p < 0.005$) with any glycemic trait (except HbA1c) in a phenome-wide rare-variant gene-level association analysis of all traits available in Common Metabolic Diseases Knowledge Portal (CMD-KP) as of July 22, 2022 (updated information may be obtained from CMD-KP links provided in Table S10)
**Condition 2:** gene is associated with fasting glucose ($p < 0.005$) in ARIC
**Condition 3:** gene is associated ($p < 0.005$) with HbA1c in a phenome-wide rare-variant gene-level association analysis of all traits available in CMD-KP as of July 22, 2022
**Condition 4:** gene is associated with HbA1c ($p < 0.005$) in ARIC

**Table S10:** Summary of information on significant genes (set-based analyses of rare variants) from Common Metabolic Diseases Knowledge Portal (as of July 22, 2022).

| Gene | Information |
|---|---|
| **Mask 1** | |
| *UGDH* | Very strong (HuGE score=45) genetic support for involvement in mean corpuscular volume (MCV)<br>Moderate (HuGE score=8.54) genetic support for involvement in 2-hour insulin<br>2-hour insulin rare-variant gene-based analysis $p = 0.007$<br>Involved in starch and sucrose metabolism; pentose and glucuronate interconversions<br>https://md.hugeamp.org/gene.html?gene=UGDH |
| *TXNDC5* | Very strong (HuGE score=45) genetic support for involvement in hemoglobin concentration (HB)<br>Important role in the regulation of iron metabolism<br>https://md.hugeamp.org/gene.html?gene=TXNDC5 |
| *ARHGEF39* | Moderate (HuGE score=3) genetic support for involvement in white blood cell count<br>Gene function not found; https://md.hugeamp.org/gene.html?gene=ARHGEF39 |
| *C15orf40* | Not much information or evidence of association with any group of metabolic traits<br>Gene function not found; https://md.hugeamp.org/gene.html?gene=C15orf40 |
| *ZNF208* | Very strong (HuGE score=45.9) genetic support for involvement in systolic blood pressure<br>Extreme (HuGE score=348) genetic support for involvement in fasting insulin adjusted for BMI<br>Significant rare-variant gene-level association of fasting insulin adjusted for BMI ($p = 1.4 \times 10^{-6}$) and HOMA-B ($p = 0.004$)<br>May be involved in transcriptional regulation; https://md.hugeamp.org/gene.html?gene=ZNF208 |
| **Mask 2** | |
| *FOSL2* | Very strong (HuGE score=45) genetic support for involvement in white blood cell count, systolic blood pressure<br>Extreme (HuGE score=152.6) genetic support for involvement in triglycerides<br>Moderate (HuGE score=4) genetic support for involvement in 2-hour insulin<br>https://md.hugeamp.org/gene.html?gene=FOSL2 |
| *RNF103-CHMP3* | Very strong (HuGE score=45) genetic support for involvement in BMI, total cholesterol, MCV<br>*RNF103* involved in protein metabolism; https://md.hugeamp.org/gene.html?gene=RNF103<br>https://md.hugeamp.org/gene.html?gene=CHMP3 |
| **Mask 3 (also Mask 4)** | |
| *CD1D* | Very strong (HuGE score=45) genetic support for involvement in mean corpuscular hemoglobin concentration (MCHC)<br>Antigen presenting protein; https://md.hugeamp.org/gene.html?gene=CD1D |
| *EGFL7* | Very strong (HuGE score=45) genetic support for involvement in lymphocyte count<br>Moderate (HuGE score=3.3) genetic support for involvement in glycemic trait adiponectin<br>Significant common variant gene-level association with MCV<br>Regulates vascular tubulogenesis in vivo; https://md.hugeamp.org/gene.html?gene=EGFL7 |
| *MIR126* | No information in CMD-KP; https://md.hugeamp.org/gene.html?gene=MIR126 |
| *AGPAT2* | Significant common variant gene-level association with red blood cell (RBC) count<br>HbA1c rare-variant gene-based analysis $p = 0.003$<br>Converts LPA to PA; https://md.hugeamp.org/gene.html?gene=AGPAT2 |
